# Validation of a New Family Values Scale Among Older Chinese Adults

**DOI:** 10.1101/2025.05.01.25326732

**Authors:** Ziyue Wang, Nadia Sourial, Xiang Zou, Howard Bergman, Xiaoyun Liu, Isabelle Vedel

## Abstract

**Objective:** To examine the reliability, validity, and factor structure of the Family Values Scale among older Chinese adults.

**Background:** Family values shape caregiving practices, intergenerational relationships, and health outcomes in later life. In China, rapid social change has transformed traditional filial norms, yet few validated instruments capture contemporary family values among older adults.

**Method:** We applied Dima’s six-step psychometric validation protocol using data from the 2018 China Longitudinal Ageing Social Survey (CLASS; N = 11,418). Exploratory and confirmatory factor analyses assessed dimensionality, while internal consistency, measurement invariance, and sensitivity analyses evaluated reliability and robustness.

**Results:** After removing two poorly performing items, the revised six-item scale demonstrated good internal consistency (Cronbach’s α = 0.80) and strong model fit in confirmatory factor analysis (CFI = 0.98; TLI = 0.96; RMSEA = 0.06). All factor loadings exceeded recommended thresholds, supporting a unidimensional structure that was invariant across key sociodemographic groups.

**Conclusion:** The six-item Family Values Scale is a reliable and valid instrument for measuring contemporary family values among older Chinese adults.

**Implications:** This parsimonious scale can be used in large population studies and policy evaluations to better understand family-based care and ageing in rapidly changing societies.

## Introduction

The unprecedented global trend in population ageing has provoked family care crises for older adults (Steinmüller, 2023; World Health Organization., 2023). In this context, family caregiving and family values - defined as “ideas about how relationships of blood, marriage, sex, and residence should relate and articulate with processes of social reproduction” (Creed, 2000) - that shape attitudes and practices in caring for older people, have gained increasing attention. Several studies have already shown that higher levels of family values are associated with better caregiving (Falzarano et al., 2021) and better health or well-being for older adults (Mao, 2015; Valdivieso-Mora et al., 2016). Furthermore, family values may also determine not merely how individual families perceive their roles in family caregiving, but also their rights, responsibilities, and ways of engageing with social institutions, the state, and the market (Steinmüller, 2023). Therefore, family values are not only as guiding norms of family members, but also have broad implications for social policy and development, yet in-depth discussions on the role of family values in caregiving from a broader societal perspective remain limited.

This gap is particularly consequential for the field of family studies, where understanding family values is fundamental to explaining intergenerational relationships, caregiving dynamics, and family well-being (Bengtson & Roberts, 1991; Silverstein et al., 2002). As societies undergo rapid demographic transitions – declining fertility rates, rising life expectancy, and increasing geographic mobility – the values that bind family members together are being renegotiated in ways that directly affect the everyday lives of older adults and their families (Daatland & Herlofson, 2005; Lowenstein, 2007). In China, where population ageing is occurring at an unprecedented pace alongside dramatic social and economic transformations (Feng et al., 2020), these shifts have created urgent, real-world challenges: who will care for the growing number of older adults, how will families negotiate evolving expectations of support and obligation, and what role should the state play in supplementing family-based care (Feng et al., 2012; Guo et al., 2022)? A validated measurement tool for contemporary family values is therefore essential not only for advancing scholarly understanding of family processes, but also for informing policy responses to the care crisis facing millions of Chinese families.

One of the reasons for the lack of in-depth research on family values is that measuring family values is challenging. First, different cultural and ethnic groups tend to use different terms to describe their family values (e.g., familism among Latinos (Cahill et al., 2021), communalism among African Americans (Schwartz et al., 2010), and filial piety among East Asians (Badanta et al., 2022)), which makes the literature on family values highly fragmented. However, researchers have also shown that these different types of family values are all rooted in the collectivist belief and share several key characteristics in common, such as maintaining high levels of family connectedness, making sacrifices for family obligations, and respecting family hierarchy (Choi et al., 2021; Christophe & Stein, 2022; Schwartz et al., 2010). These findings suggest that the tools for measuring family values in one specific context could inspire the development of measurement tools in other settings. Another challenge is the evolving and dynamic nature of family values. The process of social change has significantly changed the meaning of family in many countries. For example, in China, the focus of family life has shifted from the ancestors to the youngest generation (known as “Chinese neo-familism”)(Yan, 2016, 2018). The core of the new family interaction is to ensure the success of the child through a good education, which requires the dedication of the whole family (Yan, 2018). Nevertheless, most family value scales for Chinese populations failed to capture this new change and continue to rely on the old, static theories of parent-centred filial piety (Fu et al., 2020; Lum et al., 2016; Yeh & Bedford, 2003).

By reviewing studies on family values or filial piety scales among Chinese populations (Fu et al., 2020; Lum et al., 2016; Yeh & Bedford, 2003), we found three limitations in existing measurement tools: First, few studies have positioned their work within a broader multicultural research context. Second, most studies have not accounted for recent shifts in Chinese family values. Third, most scales were not designed for older populations. To address these gaps, this study aims to test the reliability, validity, and factor structure of the Family Values Scale, a new scale for measuring contemporary family values among older Chinese adults, in a large, nationally representative, community-dwelling sample - the China Longitudinal Ageing Social Survey (CLASS).

## Methods

This review adapted Dima’s 6-step protocol of Psychometric analyses (Dima, 2018). All materials were coded and analyzed using R 4.3.1 (R Core Team, 2023).

### Data

This study used data from the 2018 (wave 3) follow-up surveys of the China Longitudinal Ageing Social Survey (CLASS). CLASS is a nationally representative, biennial prospective cohort study of Chinese adults aged ≥ 60 years conducted by the (Blind for Review) since 2014 (According to the World Health Organization, individuals aged 60 years and older are considered older adults in developing countries (Kowal et al., 2012)). Using stratified multistage probability-proportional-to-size sampling, CLASS covered 28 of 31 province-level divisions of China, 134 counties/districts, and 462 villages/communities, enrolling 11,511 participants in the baseline survey in 2014. The sampling protocol is publicly available at (Blind for Review). The survey was also based on the collaboration networks of the Chinese Social Survey Network (CSSN), a part of the International Social Survey Program (Smith, 2001), which includes other social surveys such as the General Social Survey (GSS, the United States) (Marsden et al., 2020) and the China General Social Survey (CGSS, China) (Huhe, 2014). The CLASS has been also widely used for understanding the health and well-being of older Chinese adults with more than 400 publications (Feng et al., 2018; Jing et al., 2023; Wang et al., 2020).

Due to the study design, the questionnaire of each CLASS wave varied. In the past waves of the CLASS, only the 3^rd^ wave (2018) included family values modules. The 2018 wave of CLASS has 11,419 respondents. After removing one participant who was under the age of 60, we obtained a total of 11,418 valid participants in this study.

### Theoretical framework

Hispanic/Latino is the most extensively studied group in family values research. According to the most recent systematic review (Cahill et al., 2021), familism among Hispanic/Latino individuals can be measured by three key subdimensions (Table 1): **1. Family Support** is the emphasis on maintaining strong emotional bonds and providing mutual support among family members. **2. Family Obligation** captures the values and attitudes towards fulfilling family obligations (caring for younger or elder family members, providing financial assistance or shelter, and prioritizing time with family). **3. Family as Referents/Honour** is the notion that one’s actions and attitudes reflect on the family and contribute to the family’s honour. Literature has shown that each subdimension of family values has different effects on health outcomes or plays different roles among different ethnic groups (Chiang et al., 2019; Falzarano et al., 2021; Valdivieso-Mora et al., 2016).

**Table 1:**
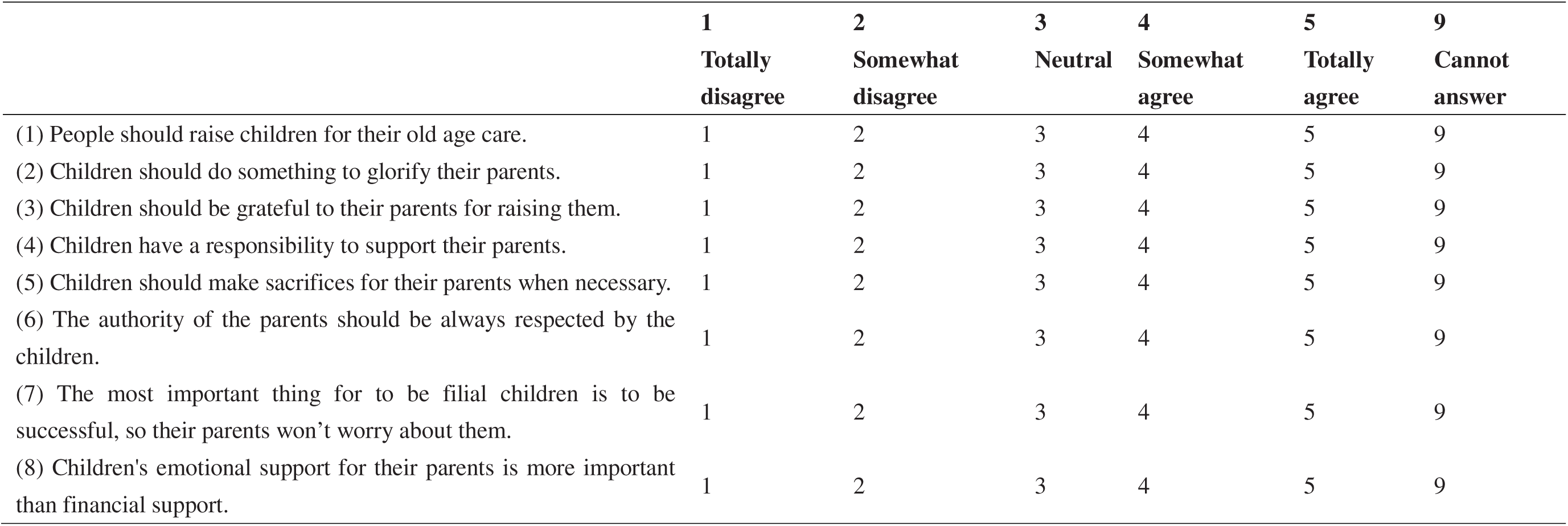
Family Value Scale (English Version) Here are some of the views on filial piety and family values, what are your thoughts on these issues?

China and other East Asian societies have long been recognized for their strong emphasis on family commitment as a core value, known as “filial piety”(He, 2021). In general terms, filial piety requires devotion to one’s parents, which is expressed through respect, support, honoring their name, having male heirs, and performing sacrifices after their death. Nevertheless, with the rise of progressivism and communism in China in the early 20th century, these traditional ideas of filial piety and family values changed significantly during the radical political, economic, and social changes in China, especially after the socialist revolution in the 1950s and the “open-door policy” in the 1980s. The pioneering scholar of Chinese family culture, Yunxiang Yan, argues that these social changes constitute the emergence of a new kind of “familism” in China (Yan called it **“Chinese neo-familism”**)(Kleinman et al., 2011; Yan, 2016, 2018, 2021). Yan highlighted several differences between Chinese neo-familism and traditional Chinese filial piety, such as the rise of the individual, junior-centered family resource allocation, and the weakening of ancestor worship. He believes that to understand China’s rapidly shifting intergenerational dynamics in contemporary China, researchers should think beyond the traditional filial piety framework and use this “Chinese neo-familism” framework.

Although several instruments have been developed to measure family values or filial piety in the Chinese context (Christophe & Stein, 2022; Fuligni et al., 1999; Steidel & Contreras, 2003; Yeh & Bedford, 2003) and some of them have been included in multiple large-scale health surveys (e.g., the Chinese Longitudinal Healthy Longevity Survey, the Chinese General Social Survey, the China Longitudinal Ageing Social Survey) (Center for Healthy Ageing and Development Study, 2002; National Survey Research Centre, 2016; Zhang & Xiang, 2021), the structure of family values in China remains unclear, particularly within the new context of “Chinese neo-familism”. By validating the Family Values Scales in this study, we aim to identify the factor structures of family values among older Chinese adults and test the two relevant frameworks: the three-dimensional framework proposed by Cahill et al. versus the unified “neo-familism” framework from Yan.

### Measurements: Family Values Scale

The Family Values Scale is an 8-item scale that aims to evaluate respondents’ agreement with family values views (See the full scale in Table 1), including family relationships/intimacy (e.g., “Children should be grateful to their parents for upbringing.”), family responsibility (e.g., “Children have a responsibility for supporting their parents.”), and power/authority in the family (e.g., “The authority of the parents should be always respected by the children.”). These items were rated in terms of respondents’ agreement by a 5-point Likert Scale (1 = Totally disagree, 2 = Somewhat disagree, 3 = Neutral, 4 = Somewhat agree, 5 = Totally agree), with a score of 9 assigned to respondents who could not answer. A higher score indicates a higher level of familism among the participants. The response = 9 was coded as a missing value and all missing values in the scale were imputed by multiple imputations with the predictive mean matching imputation method since the scores of items are not normally distributed (Morris et al., 2014).

### Statistics

#### Descriptive analysis

We conducted a descriptive analysis to identify any missing values, outliers, and reverse-coded items. Outliers are considered as the Mahalanobis distance statistics (D^2^ values) with probability values <.001 (Tabachnick et al., 2013). Reverse-coded items were detected using a plot of Spearman correlation coefficients. Responses of 1 or 2 (“Totally disagree” or “Somewhat disagree”) were classified as “disagree” and responses of 3-5 (“Neutral”, “Somewhat agree”, or “Totally agree”) were classified as “agree”. The distribution of responses for each item was used to detect any ceiling or floor effects (Usually, ≥ 15% of respondents achieving the lowest or highest possible score is considered a floor or ceiling effect (Terwee et al., 2007)).

#### Validity & Factor structure

We conducted both exploratory and confirmatory factor analyses to provide a complementary perspective on the dimensionality of these items. First, the Kaiser-Meyer-Olkin (KMO) Test (Threshold criteria: KMO statistic > 0.5) and Bartlett’s sphere tests (Threshold criteria: P < 0.05) were conducted to confirm the suitability of data for factor analysis. Second, the dataset was divided into two random subsets: the training set (70% observations) and the test set (30% observations). Then, the training set was further divided into two random subsets (each containing 35% observations of the original dataset) for the parallel analysis of two exploratory factor analyses (Principal Component Analysis, PCA vs. Factor Analysis, FA). The scree plots of eigenvalues of factors or principal components were used to guide the choice of the number of factors. The number of factors/components were identified at the point where the curves of scree plots start to flatten (known as the “elbow criteria” (Zoski & Jurs, 1996)). Finally, the results of the exploratory factor analyses were further tested in the test set using confirmatory factor analyses, including the model fit (Tucker-Lewis index (TLI) and Comparative Fit Index (CFI) >0.95; root mean square error of approximation (RMSEA) <0.06) and factor loadings (>.30 or .40 for the hypothesized dimensions) (Floyd & Widaman, 1995).

In addition to factor analyses, we also used other analyses to help us determine the factor structure of our scale. Very Simple Structure (VSS) analysis seeks a structural solution in which each item belongs to one main factor with the lowest loading on other factors (Revelle & Rocklin, 1979). Item cluster analysis (ICLUST) and hierarchical factor analysis can determine the factor structures by identifying correlated items or hierarchical organization (Revelle, 1979). These results were reported in Appendix A.

To examine the model invariance of the scale, we conducted multigroup analyses by sex (male vs. female), education (primary school or lower vs. middle school or higher), locality (urban vs. rural), geographical regions (Eastern, Central, and Western China), and activities of daily living (ADL dependency or not). We followed the conventions of analyses from less restrictive to the most restrictive models: configural invariance, metric invariance, and scalar invariance. As the Chi-square test for the goodness of fit is sensitive to sample size (Bentler & Bonett, 1980), we used the criterion of a 0.01 change in CFI and a 0.015 change in RMSEA to determine model fit (Putnick & Bornstein, 2016).

#### Reliability

We used several indices to test the reliability of the scale and its sub-scales (if applicable), including Cronbach’s alpha, Guttman’s lambda 6, beta, and omega (Dunn et al., 2014). We also tested the scale properties if any items were dropped to see if we could improve the internal consistency by removing any existing items.

#### Sensitivity analyses

To address the potential bias introduced by missing values and outliers, we repeated all analyses with two datasets: (1) Removing all outliers detected by D^2^ values in the descriptive analysis (Appendix A). (2) Removing all participants with missing values on the scales (complete case analysis (Morris et al., 2014), Appendix B). We carefully examined all the results to see if any of our findings were driven by particular data points.

## Results

### Participants Characteristics

The study sample consisted of 11□418 older Chinese adults with a mean age of 71.5 years. 50.2% of the participants were women, 69.3% were married or living with a partner, and 42.3% lived in rural areas (Table 2). Almost half of the older adults in our sample lived in Eastern China (N = 5 676, 49.7%) and less than one-third had completed middle school or higher (N = 3 746, 32.8%). Regarding health status and health service unitization, the participants had an average of 1.4 chronic diseases. 8.3% of participants had a moderate or higher level of dependence on ADL, and 27.5% of participants had one or more hospitalizations in the past year. In terms of family and social support for older people, the average number of people in their households and the average number of their children is 2.6. According to the result of the Lubben Social Network Scale (Chang et al., 2018), more than one-third of participants were at risk of social isolation (N = 3873, 33.9%)

**Table 2:**
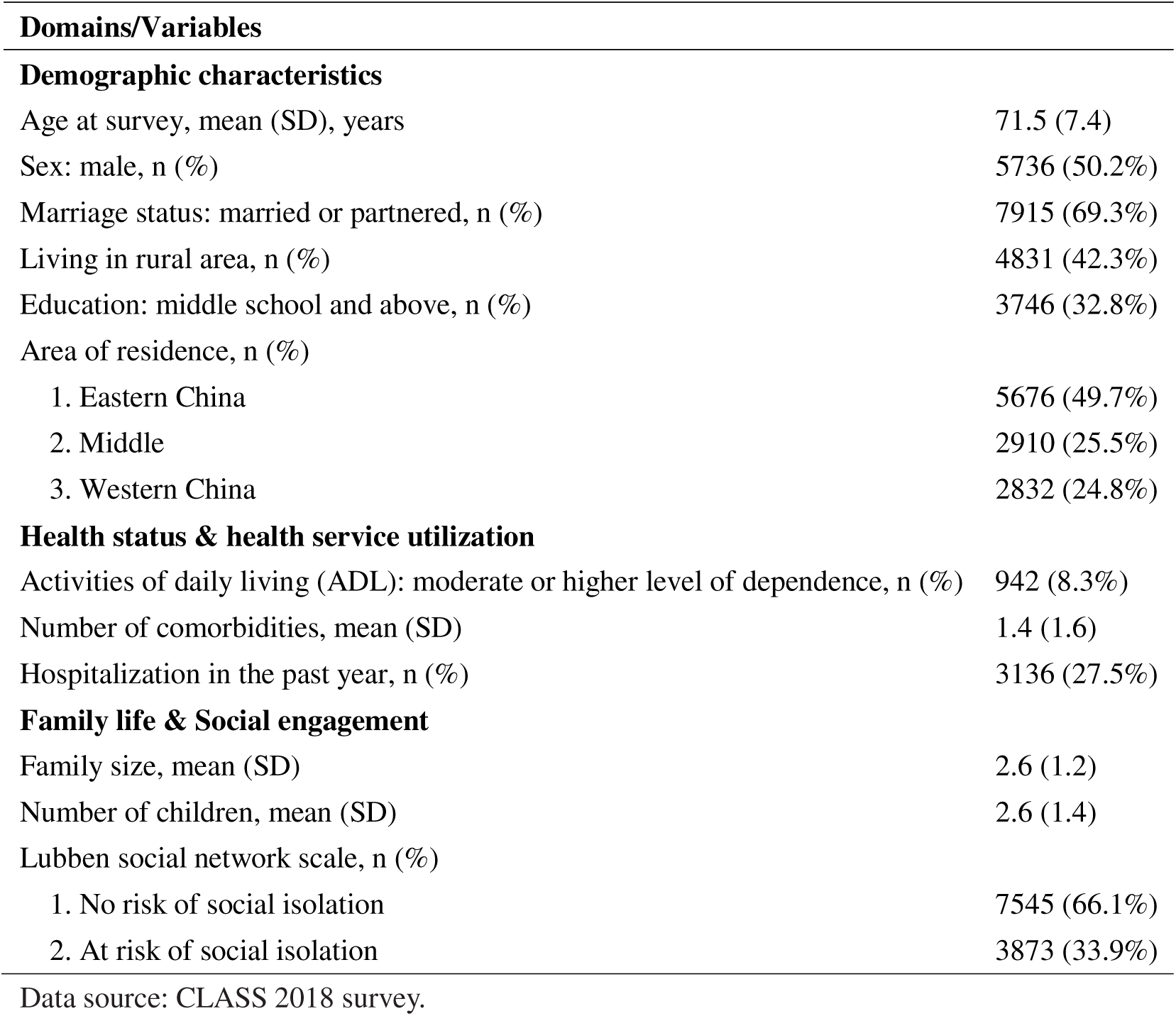
Participant characteristics (N=11418)

### Descriptive analysis of the Family Value Scale

Seven of the eight items of the Family Value Scale had less than 5% missing values (Table 3). Only one item (item 5) had more than 5% missing (5.47%) and the proportion of missing values in the total score was 7.57%. However, for each item on the scale, the agreement rate exceeded 85% among participants. Item 5 had the highest proportion of disagreement (N = 1 729, 15.14%), while the other items were all agreed by more than 90% of the participants. The mean total score of the Family Value Scale was 16.15 (standard deviation (SD): 4.48, Appendix A). The correlation matrix showed that all items were significantly and positively correlated with each other, with no evidence of reverse coding (P<0.01, Appendix C1). Although the correlation between item 1/item 5 and other items (r = 0.15-0.35) was relatively low, these correlations were still statistically significant. We identified 230 respondents as outliers (D^2^ values with probability values <.001. Maximum D^2^ value: 58.55, Appendix A).

**Table 3:** Descriptive statistics of all items.

| Items | No. | Score | | | | | 1 to 3 (%) | 4 to 5 (%) | Missing ratio (%) | Mean ± SD | Skew | Kurtosis |
| --- | --- | --- | --- | --- | --- | --- | --- | --- | --- | --- | --- | --- |
|  |  | 1 | 2 | 3 | 4 | 5 |  |  |  |  |  |  |
| Item1 | 11116 | 3732 | 4955 | 1743 | 400 | 286 | 91.35 | 6.01 | 2.72 | 1.97 ± 0.93 | 1.09 | 1.32 |
| Item2 | 10999 | 3488 | 4930 | 2047 | 433 | 101 | 91.65 | 4.68 | 3.81 | 1.98 ± 0.86 | 0.76 | 0.48 |
| Item3 | 11038 | 3430 | 5314 | 1928 | 313 | 53 | 93.47 | 3.21 | 3.44 | 1.94 ± 0.80 | 0.68 | 0.44 |
| Item4 | 11065 | 3667 | 5064 | 1918 | 376 | 40 | 93.27 | 3.64 | 3.19 | 1.92 ± 0.82 | 0.68 | 0.22 |
| Item5 | 10826 | 2472 | 4104 | 2521 | 1335 | 394 | 79.67 | 15.14 | 5.47 | 2.36 ± 1.07 | 0.55 | -0.37 |
| Item6 | 11033 | 3115 | 5199 | 2177 | 447 | 95 | 91.88 | 4.75 | 3.49 | 2.02 ± 0.85 | 0.70 | 0.46 |
| Item7 | 11013 | 3096 | 4781 | 2259 | 756 | 121 | 88.77 | 7.68 | 3.68 | 2.09 ± 0.92 | 0.67 | 0.08 |
| Item8 | 11073 | 3792 | 4899 | 2008 | 336 | 38 | 93.70 | 3.28 | 3.12 | 1.91 ± 0.82 | 0.65 | 0.10 |
| Total | 10554 | - | - | - | - | - | - | - | 7.57 | 16.15 ± 4.48 | 0.35 | -0.04 |

SD: Standard deviation.

### Factor structure

Both Kaiser-Meyer-Olkin (KMO=0.87) and Bartlett’s sphere tests (χ2 = 21197.98, df=28, P<0.001) confirmed the data were suitable for factor analysis. In the parallel analysis (Appendix C2), PCA results supported the assumption of a unidimensional structure. While the FA result suggested three factors on the scale, there was only one factor before the “elbow point” (the point before the line in the scree plot flattens out) in the curve, which also suggested a one-factor structure. The VSS results further confirmed a unidimensional interpretation as the goodness of fit was highest at one factor and decreased as the number of factors increased. The results of ICLUST and hierarchical factor analysis also supported a unidimensional scale solution (Appendix A).

To further test the unidimensional scale solution, we conducted a 1-factor CFA of the 8-item Family Value Scale. The CFA result showed a low goodness of fit (CFI = 0.94; TLI = 0.91; RMSEA = 0.08 [95% CI: 0.07–0.08]). Although the factor loadings of each item are within the expected range (>0.30) for the hypothesized one dimension, item 1 and item 5 showed the lowest factor loading (0.37 and 0.39 respectively, Appendix C3). By removing item 1 and item 5, the goodness of fit of our new 6-tem scale was significantly improved (CFI = 0.98; TLI = 0.96; RMSEA = 0.06 [95% CI: 0.05–0.07]). The 1-factor structure explained 40.77% of the variance in the 6-item Family Values Scale (Appendix A). The results of the CFA therefore confirmed the one-factor assumption in EFA.

We also tested the measurement invariance of the Family Value Scale for sex, education, urban/rural classification, geographical regions, and ADL dependency. The fit indices for each subgroup demonstrated strong measurement invariance across these categories (Appendix D).

### Reliability

The 8-item scale showed good internal consistency with Cronbach’s α, Guttman’s λ 6, and ω, but not β coefficient, which is a more conservative estimate of internal consistency (Cronbach’s α = 0.80 (95% CI: 0.78 - 0.82), Guttman’s λ 6 = 0.78, β = 0.55, and ω = 0.79 (95% CI: 0.78 - 0.80)). After removing item 1 and item 5, the result of internal consistency tests showed that all reliability indices of the Family Value Scale (Cronbach’s α = 0.80 (95% CI: 0.79 - 0.81), Guttman’s λ 6 = 0.78, β = 0.75, and ω = 0.80 (95% CI: 0.79 - 0.81)) were within the acceptable ranges. We found no evidence that removing additional items would improve the scale’s internal consistency, confirming that the 6-item Family Values Scale is the best measurement (Table 4).

**Table 4:**
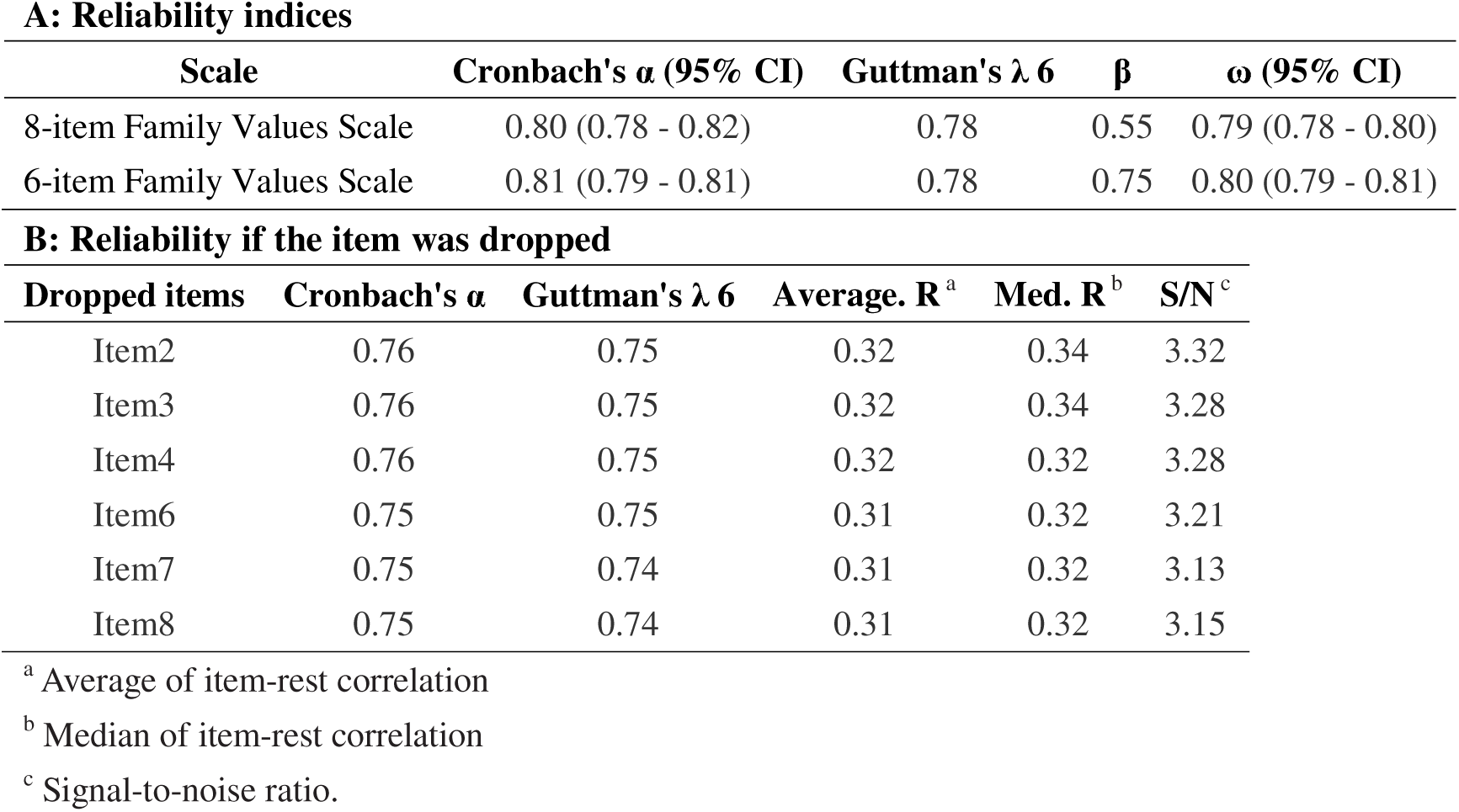
Results of the internal consistency test.

**A: Reliability indices**
| Scale | Cronbach's $\alpha$ (95% CI) | Guttman's $\lambda$ 6 | $\beta$ | $\omega$ (95% CI) |
| --- | --- | --- | --- | --- |
| 8-item Family Values Scale | 0.80 (0.78 - 0.82) | 0.78 | 0.55 | 0.79 (0.78 - 0.80) |
| 6-item Family Values Scale | 0.81 (0.79 - 0.81) | 0.78 | 0.75 | 0.80 (0.79 - 0.81) |

**B: Reliability if the item was dropped**
| Dropped items | Cronbach's $\alpha$ | Guttman's $\lambda$ 6 | Average. R<sup>a</sup> | Med. R<sup>b</sup> | S/N<sup>c</sup> |
| --- | --- | --- | --- | --- | --- |
| Item2 | 0.76 | 0.75 | 0.32 | 0.34 | 3.32 |
| Item3 | 0.76 | 0.75 | 0.32 | 0.34 | 3.28 |
| Item4 | 0.76 | 0.75 | 0.32 | 0.32 | 3.28 |
| Item6 | 0.75 | 0.75 | 0.31 | 0.32 | 3.21 |
| Item7 | 0.75 | 0.74 | 0.31 | 0.32 | 3.13 |
| Item8 | 0.75 | 0.74 | 0.31 | 0.32 | 3.15 |

<sup>a</sup> Average of item-rest correlation<sup>b</sup> Median of item-rest correlation<sup>c</sup> Signal-to-noise ratio.

### Sensitivity analyses

The analysis results excluding outliers are presented in Appendix A, and the complete case analysis is shown in Appendix B. The findings from our main analysis closely aligned with these additional analyses.

## Discussion

To the best of our knowledge, this is the first study that tested the validity of a family value-related scale, including the reliability, validity, and factor structure, in a large nationally representative, community-dwelling sample of older Chinese adults. Our results confirmed that the Family Values Scale with a unidimensional factor structure has good reliability and validity in measuring family values among older Chinese adults.

In contrast to numerous studies of other ethnic groups (e.g., Hispanics/Latinos, African Americans) (Chiang et al., 2019; Travers et al., 2020; Valdivieso-Mora et al., 2016), our analyses support a one-factor solution of the Family Values Scale with no evidence of sub-dimensional structures. This unique finding probably reflects the changing nature of family relationships in China today. For example, item 5 (“Children should make sacrifices for their parents when necessary.”) is related to family obligations or subjugations, and Item 1 (“People should raise children for their old age care.”) is a very traditional filial piety practice in China (Yeh & Bedford, 2003). Both of them can be found in many scales of family values or filial piety (Christophe & Stein, 2022; Fuligni et al., 1999; Steidel & Contreras, 2003; Yeh & Bedford, 2003). However, neither of these two items shows a good fit for our scale, and item 5 even has the most disagreements in our scale. Item 1 and item 5 also have significantly lower correlation coefficients (<0.30) of other items. By removing item 5 and item 1, the goodness of fit of the scale was significantly improved. As Yan and other researchers suggested, the core of the Chinese new family practice is to ensure the success of the child (usually through a good education) (Yan, 2021). Many traditional values of filial piety, especially authoritarian or obligation aspects such as obedience to one’s parents, fulfilling one’s parents’ expectations, and being respectful to ancestors, are much less prevalent in today’s Chinese family interactions (Yan, 2016, 2018). In our scale, item 7 (“The most important thing for to be filial children is to be successful, so their parents won’t worry about them”) reflected this new intergenerational dynamic and it showed good reliability and validity.

This study has several strengths. First, our dataset is a large, nationally representative sample of older Chinese adults with more than eleven thousand participants (N=11 418). To our knowledge, this is the largest study of family values validation among older Chinese adults and the largest of its kind in the world. Second, our scale considers items that reflect contemporary Chinese familism, which allows it to capture the recent changes in family values among older Chinese adults. Third, our 6-item scale is the shortest scale of existing filial piety or family values scales for older Chinese adults, suggesting its good applicability in large-scale research. Our study also has limitations. First, the sample was representative of community-based older people in China. The scale may not apply to other research settings (e.g., older adults in high-income countries, or older Chinese people in long-term care facilities) and the results of these external measurements should be interpreted with caution. Second, the high agreement rates suggested a low variability of responses and the existence of the ceiling effect, which also raised a concern that our scale may lack the capacity to differentiate between individuals with different attitudes toward the measured construct. This concern limits the ability of this scale to capture nuanced differences in family values.

### Implications

Our findings carry several important implications for family studies and social policy. First, the validated Family Values Scale provides family researchers with a reliable, parsimonious instrument that can be readily incorporated into large-scale surveys and longitudinal studies examining how family values shape caregiving arrangements, intergenerational solidarity, and family well-being among ageing populations (Bengtson & Roberts, 1991; Silverstein et al., 2002). Second, the unidimensional structure of the scale offers empirical support for Yan’s “Chinese neo-familism” framework (Yan, 2018, 2021), suggesting that contemporary Chinese family values have converged into a unified orientation that transcends the traditional distinctions between support, obligation, and honour. This finding has direct relevance for family scholars studying how modernization and social change reshape family relationships across diverse cultural contexts (Daatland & Herlofson, 2005; Lowenstein, 2007). Third, from a social policy perspective, the scale can serve as a practical tool for evaluating the effectiveness of elder care policies and family support programs. As China’s ageing population continues to grow rapidly (Feng et al., 2020), policymakers face the pressing challenge of designing interventions that align with evolving family values rather than relying on assumptions rooted in traditional filial piety (Feng et al., 2012). By measuring contemporary family values at the population level, researchers and policymakers can better identify communities where family-based care systems are under strain and where formal support services are most urgently needed (Guo et al., 2022). Finally, the cross-cultural measurement approach adopted in this study – grounding the Chinese scale within a broader multicultural framework of familism – opens pathways for comparative family research across societies experiencing similar demographic and cultural transitions (Lowenstein, 2007), contributing to a more globally informed understanding of family processes in later life. In conclusion, our validation of the Family Values Scale confirmed that this scale has good reliability and validity in capturing family values among older Chinese adults. The unidimensional factor structure of this scale also showed adequate goodness of fit and reflected the contemporary family values among Chinese people. This new Family Values Scale provided a useful tool for rapidly measuring current family values in large community-based samples of older Chinese adults. This new tool could be useful for future researchers in family caregiving for older adults and for policymakers to evaluate social policies in elderly care. Other methodologies of validation of interpretation of the Family Values Scales may also help researchers from other countries to measure the concept of family values in their own research settings.

## Data Availability

The data that support the findings of this study are available from the National Survey Research Center at Renmin University of China. Restrictions apply to the availability of these data, which were used under license for this study. Data are available at http://class.ruc.edu.cn/ with the permission of the National Survey Research Center at Renmin University of China.

## ACKNOWLEDGMENTS

We thank Dr. Tovah Cowan (Douglas Mental Health University Institute, Montreal, Canada) and Joon Lee (The Research Institute of the McGill University Health Centre, Montreal, Canada) for their assistance during this study. We thank Dr. Rize Jing at Renmin University of China for his guidance on the China Longitudinal Aging Social Survey dataset. We also thank Gabriela Lopes (McGill University) for editing the manuscript.

## ETHICAL APPROVAL

Since this study was based on publicly available data, additional ethical approval or informed consent was not required.

