## Appendix A for "Validation of a New Family Values Scale Among Older Chinese Adults"

**Appendix A: Validation Analyses with Outliers Removed**

**Table A1. Distribution of item response options**

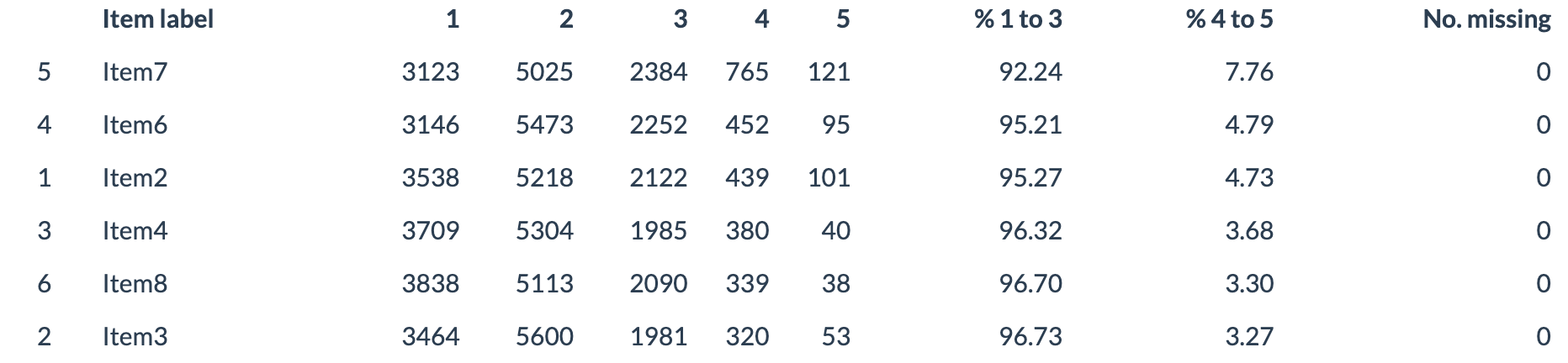

**Table A2. Descriptive statistics for individual items**

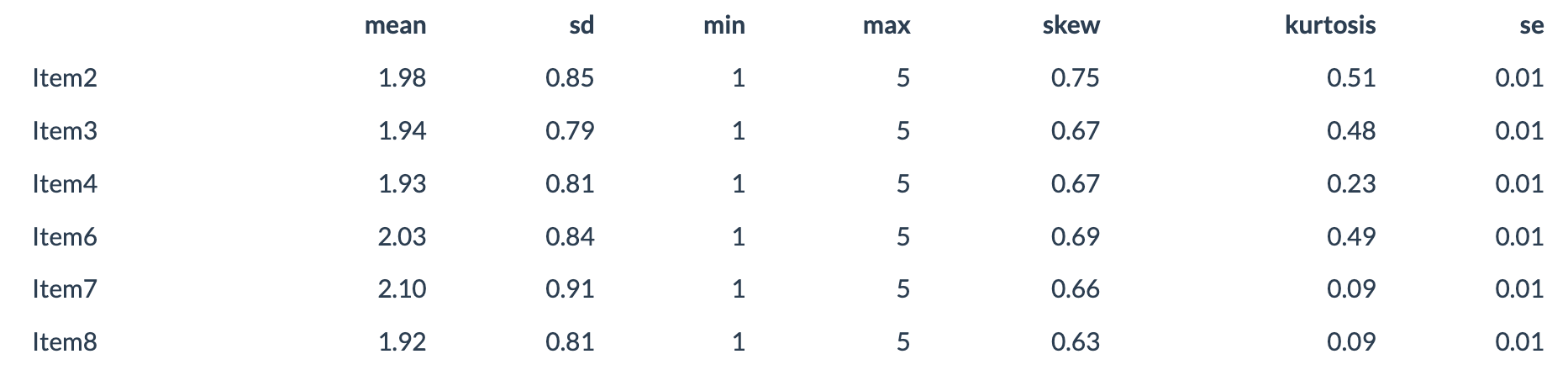

**Table A3. Reliability indices for all scales**

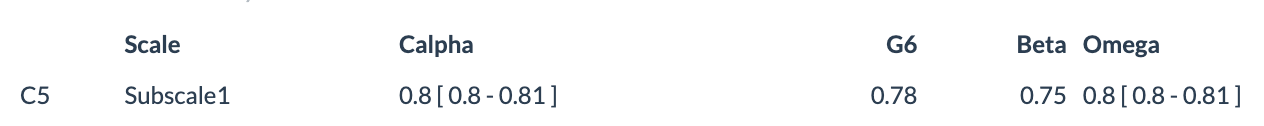

**Table A4. Reliability coefficients with each item removed**

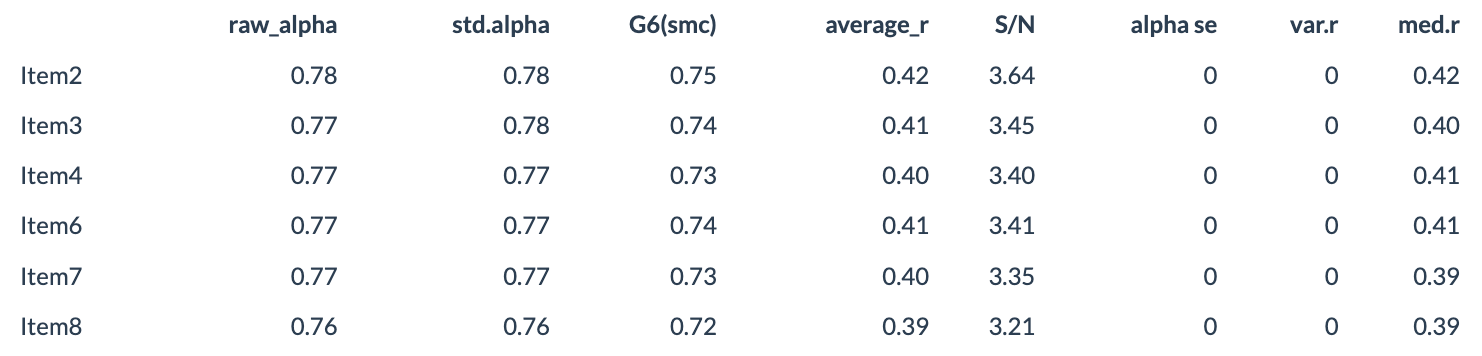

**Table A5: Descriptive statistics for total scale scores**

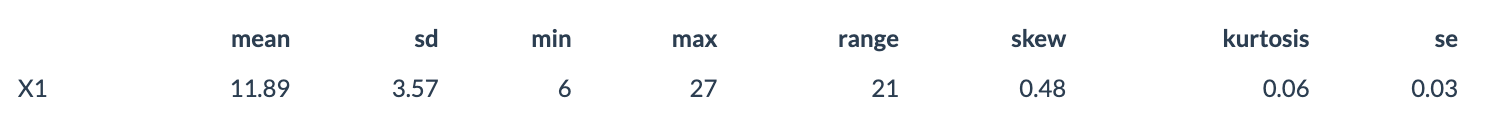

**Figure A1: Bar plots of high score response frequencies**

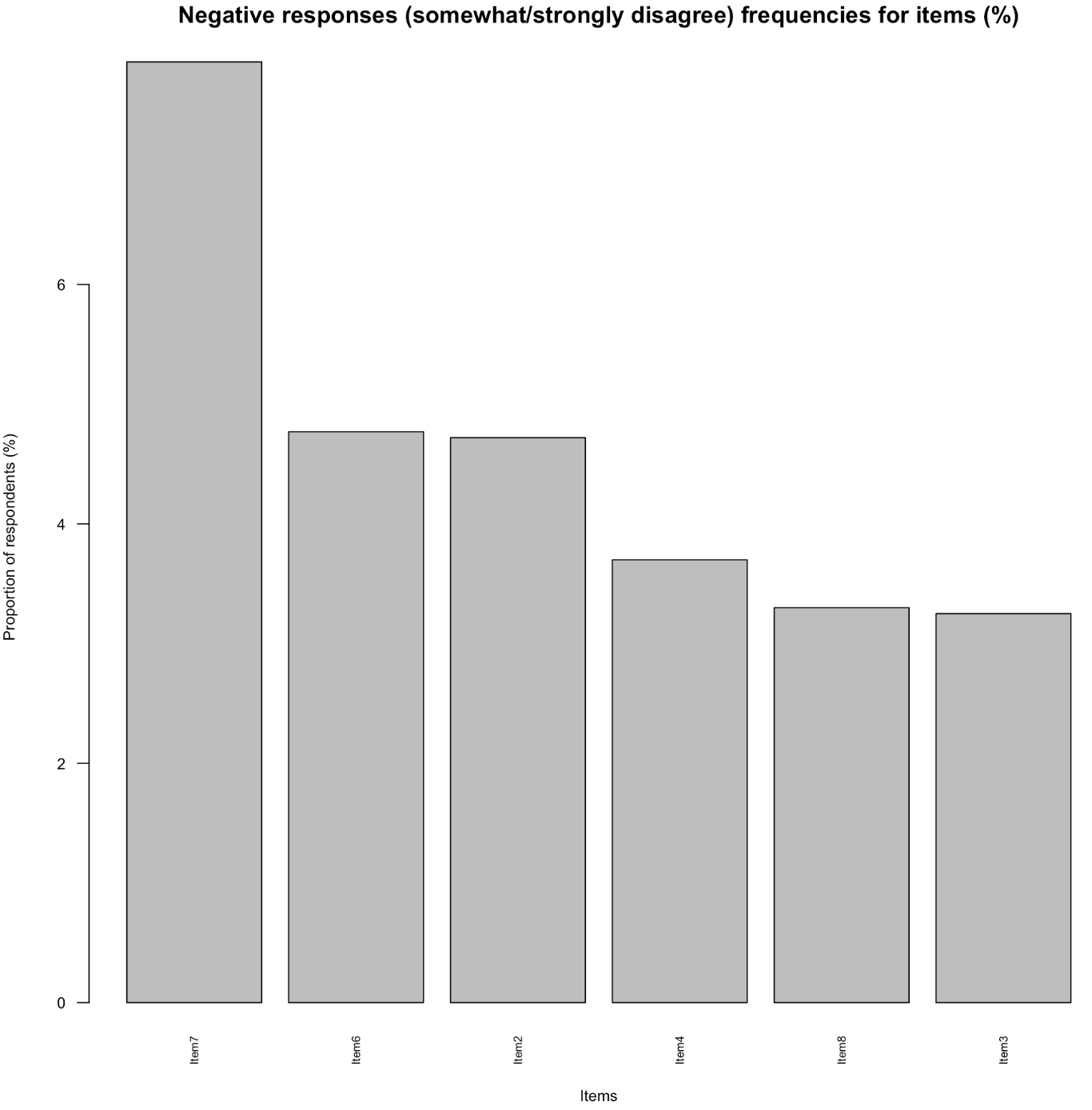

**Figure A2: Bar plots of response frequencies**

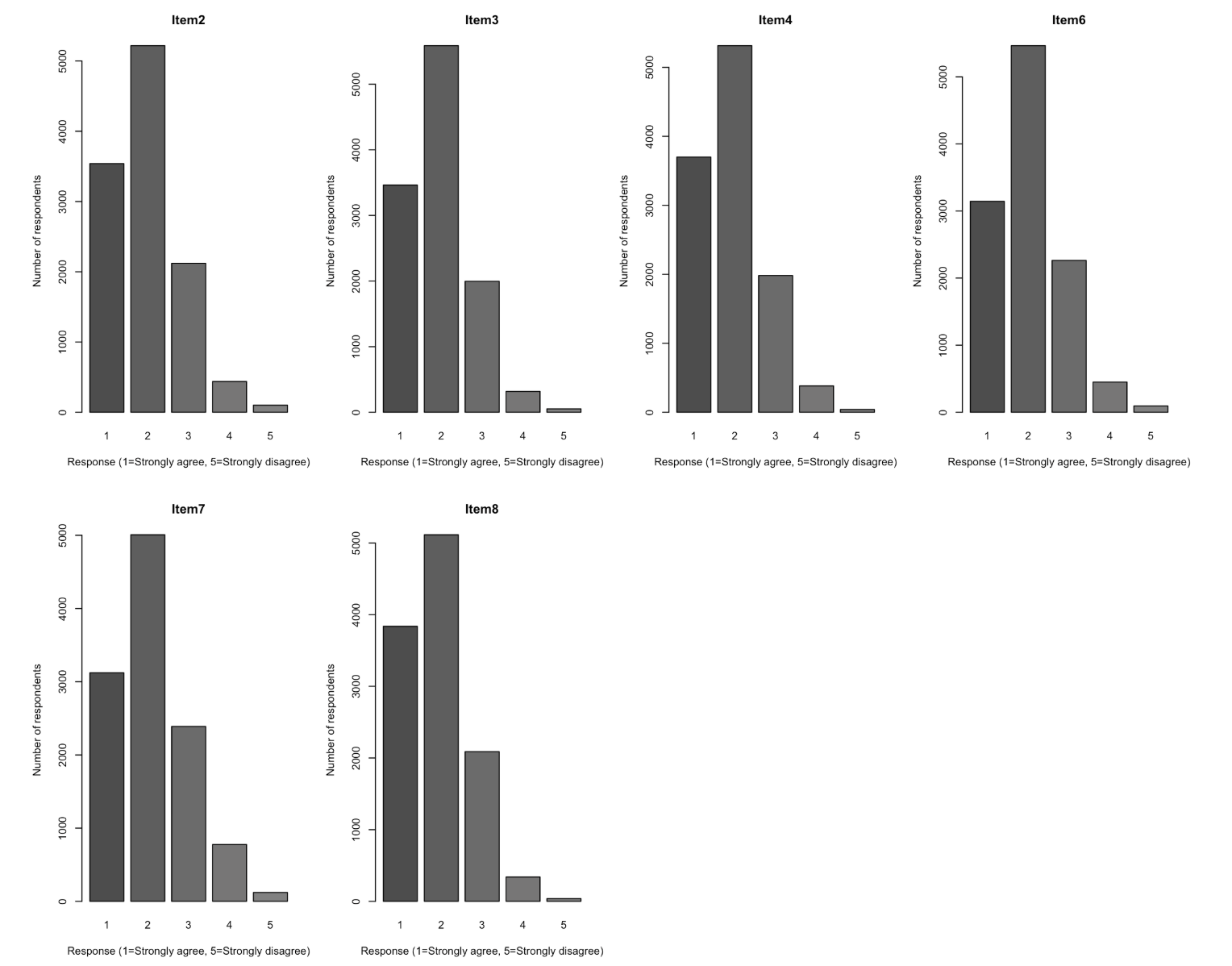

**Figure A3: Heatmap of Spearman correlations among item scores**

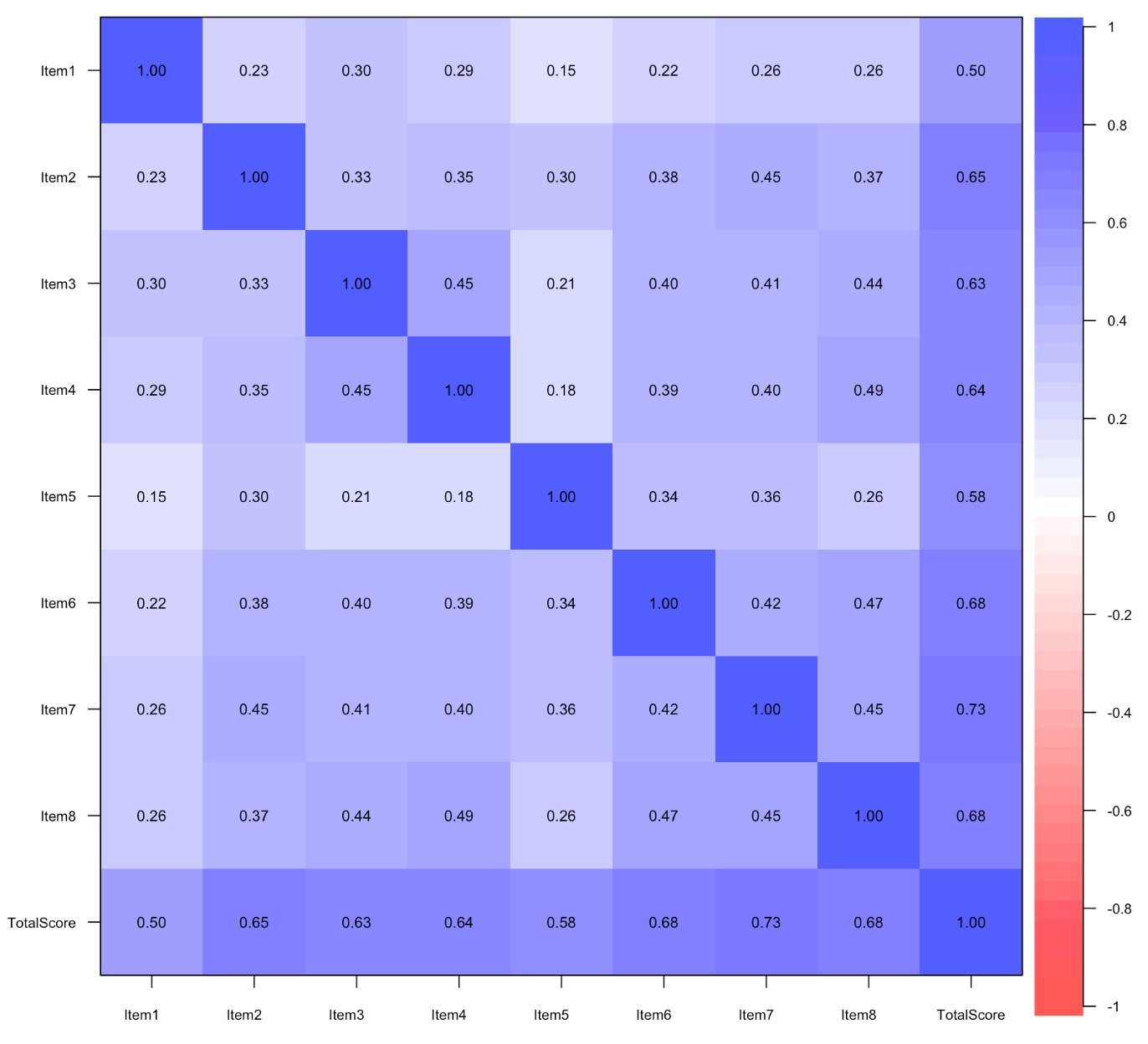

**Figure A4: Identification of multivariate outliers in item set (Q–Q plot)**

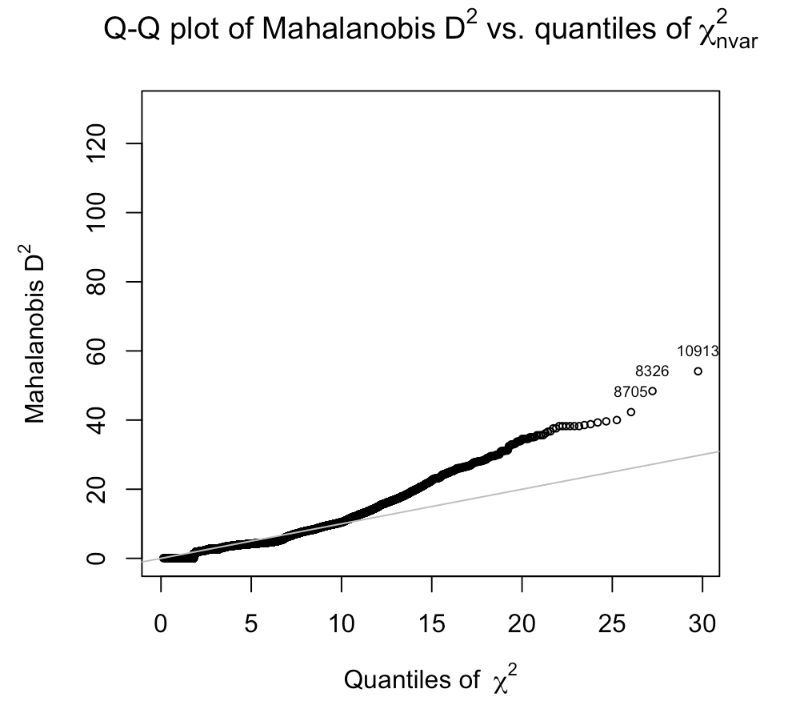

**Figure A5: Identification of multivariate outliers in item set (histogram)**

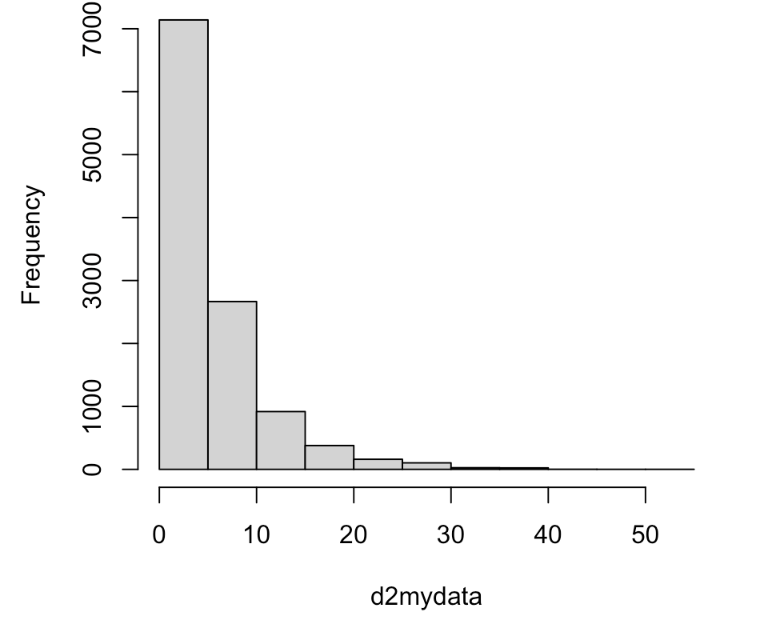

**Figure A6: Parallel analysis scree plot for factor extraction**

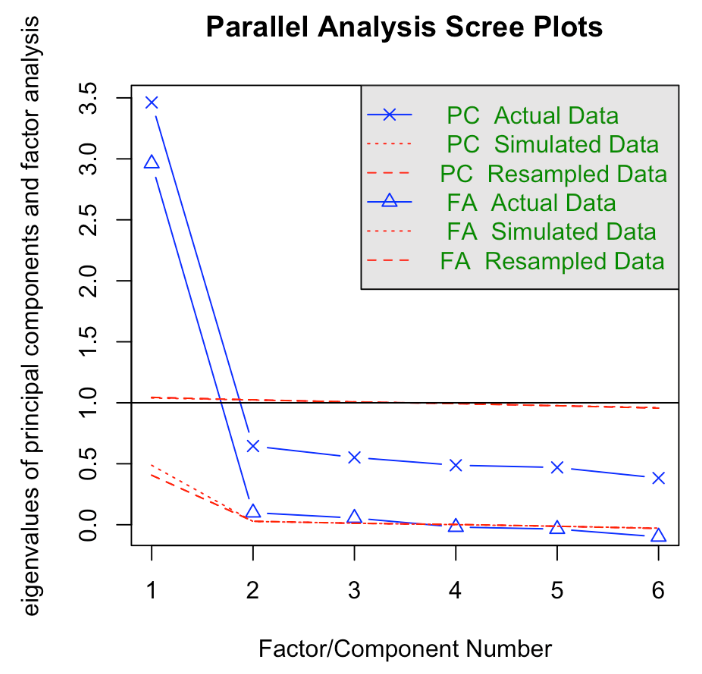

**Figure A7: Very Simple Structure (VSS) plot including outliers**

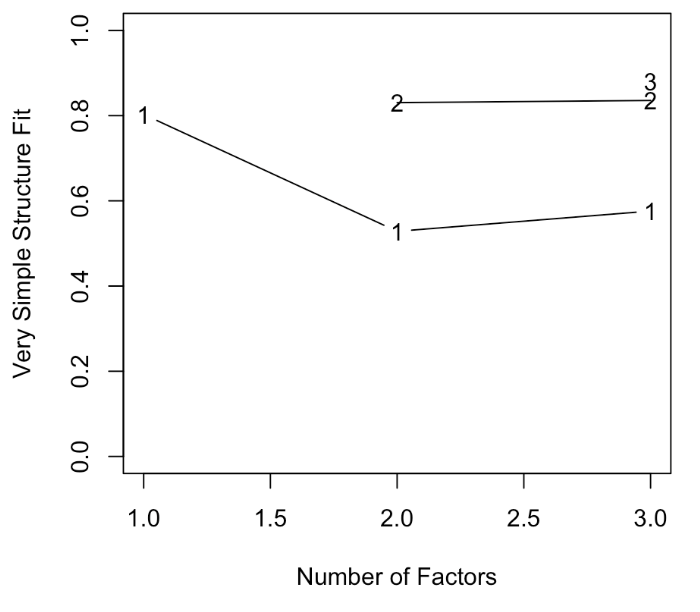

**Figure A8: Very Simple Structure (VSS) plot excluding outliers**

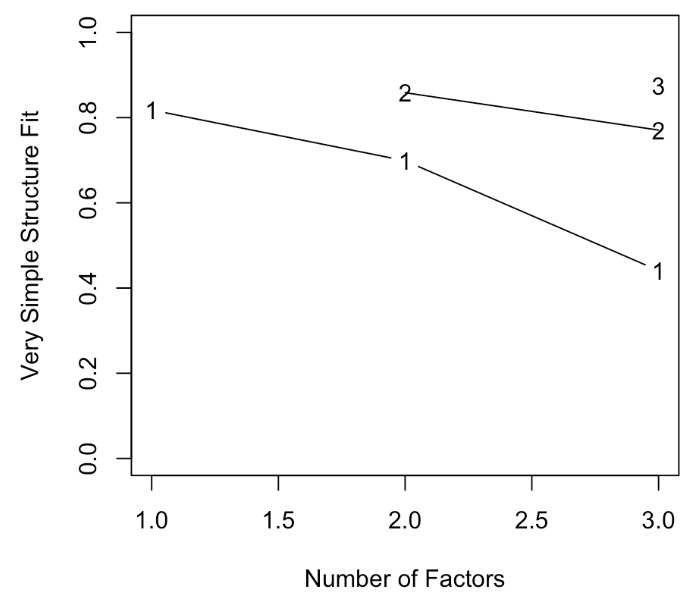

**Figure A9: One-factor exploratory factor analysis (principal axis factoring), including outliers**

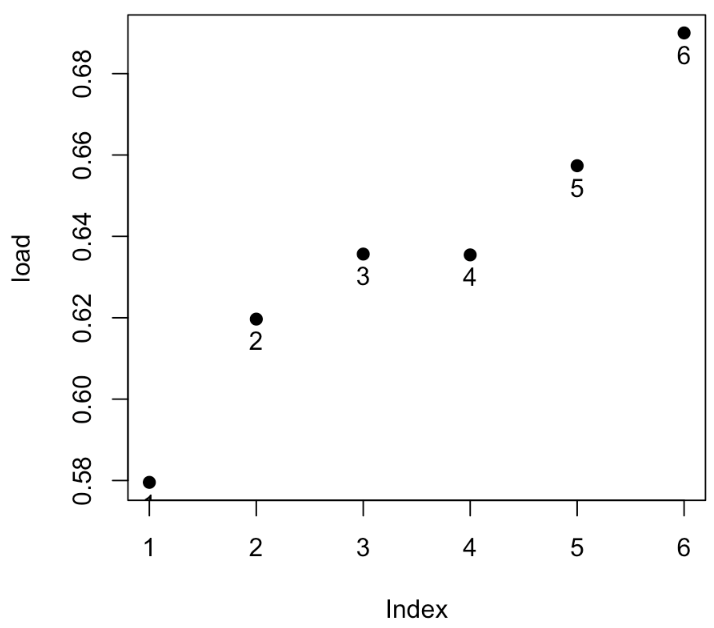

**Figure A10: One-factor exploratory factor analysis (principal axis factoring), excluding outliers**

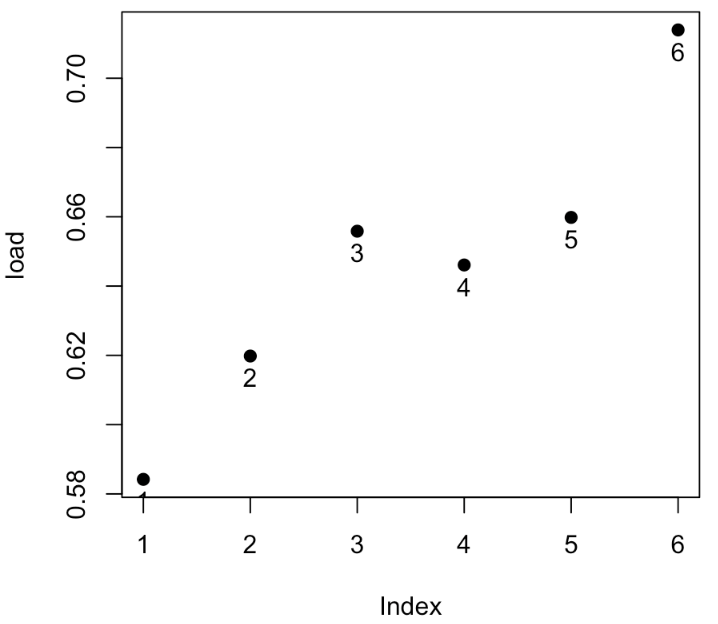

**Figure A11: Hierarchical cluster analysis of scale items (ICLUST), including outliers**

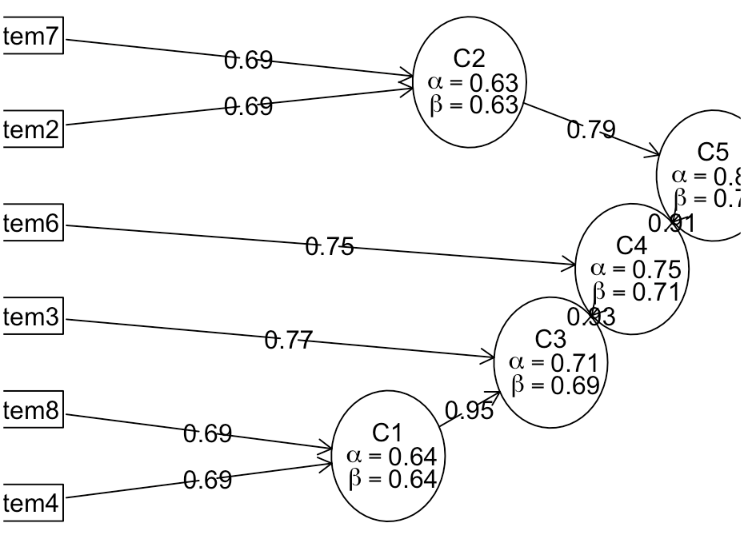

**Figure A12: Hierarchical cluster analysis of scale items (ICLUST), excluding outliers**

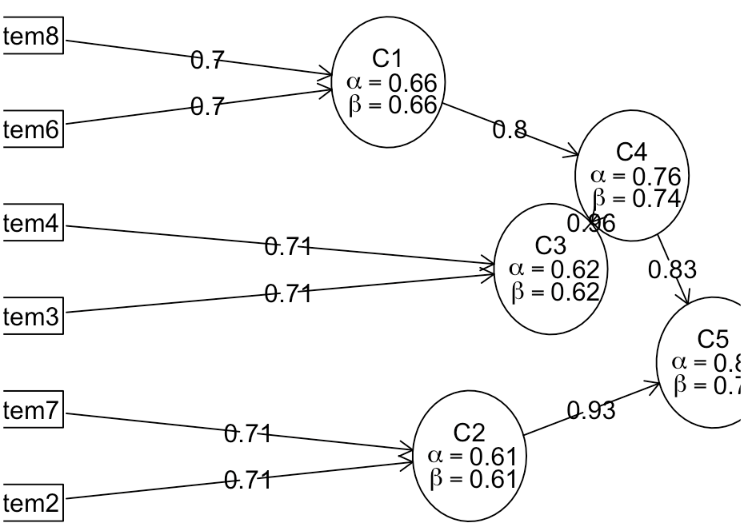

**Figure A13: One-factor confirmatory factor analysis model, including outliers**

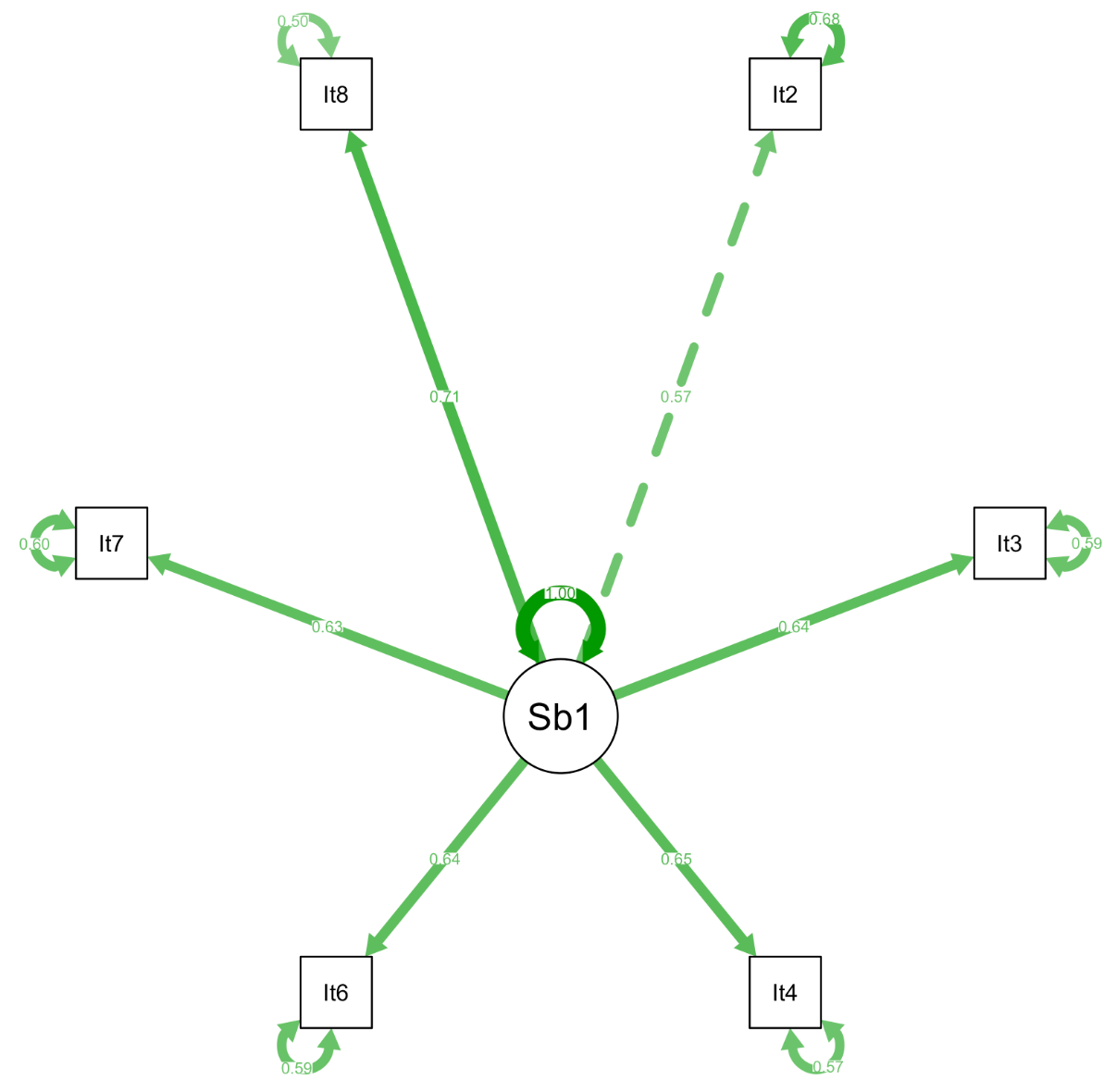

**Figure A14: One-factor confirmatory factor analysis model, excluding outliers**

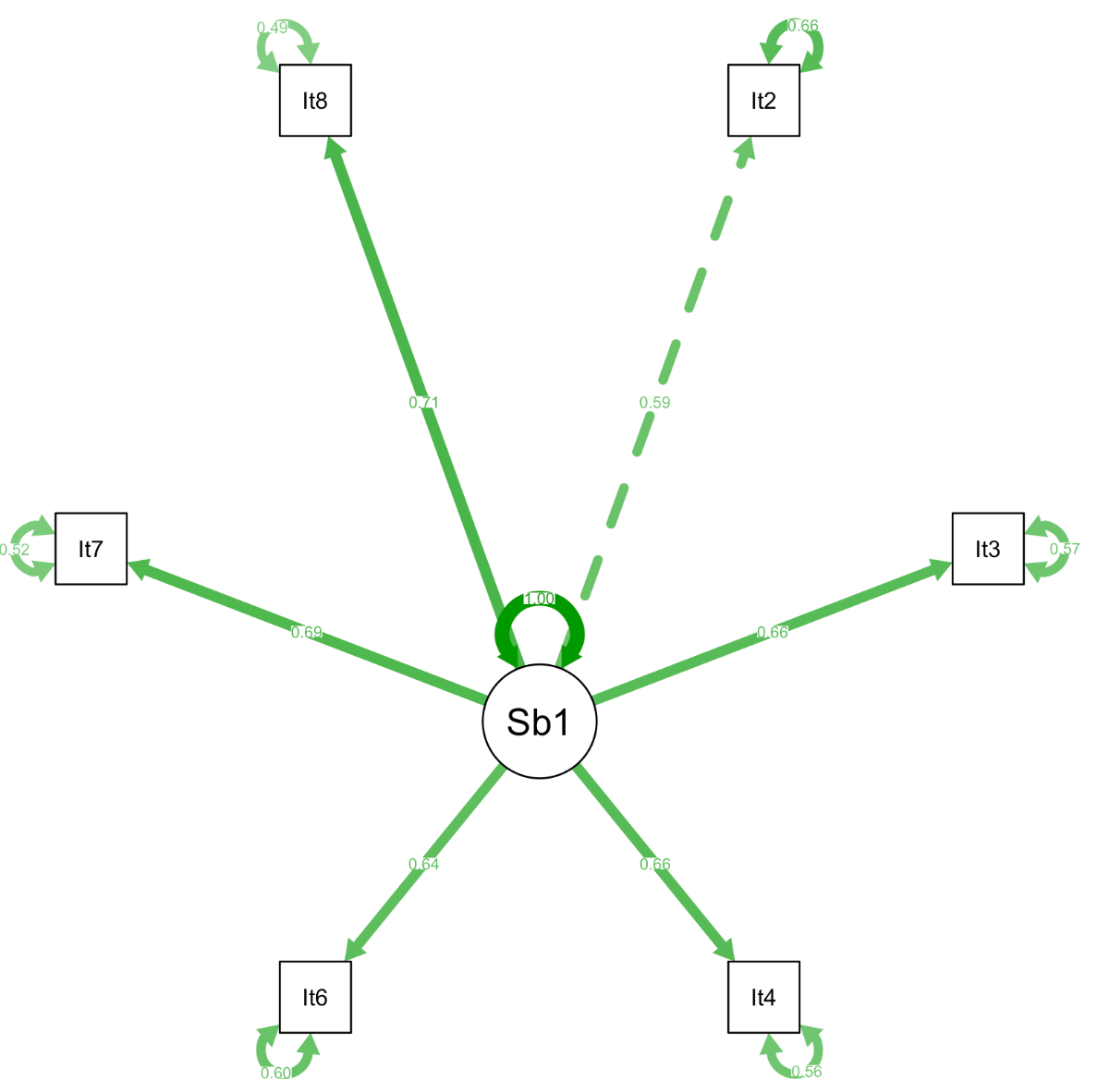

**Figure A15: Distribution of total scale scores**

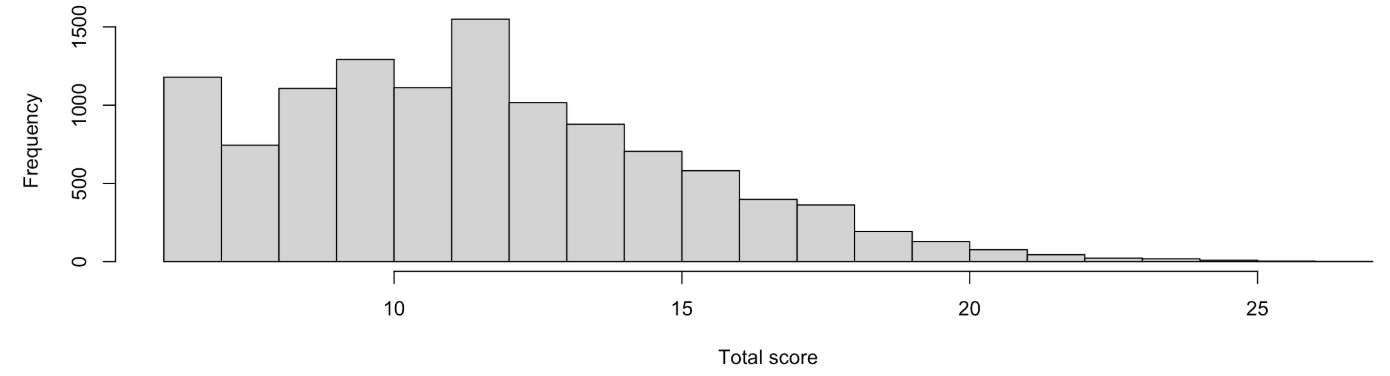
