## Appendix B for "Validation of a New Family Values Scale Among Older Chinese Adults"

**Appendix B: Validation Analyses with Outliers Removed**

**Table B1. Distribution of item response options**

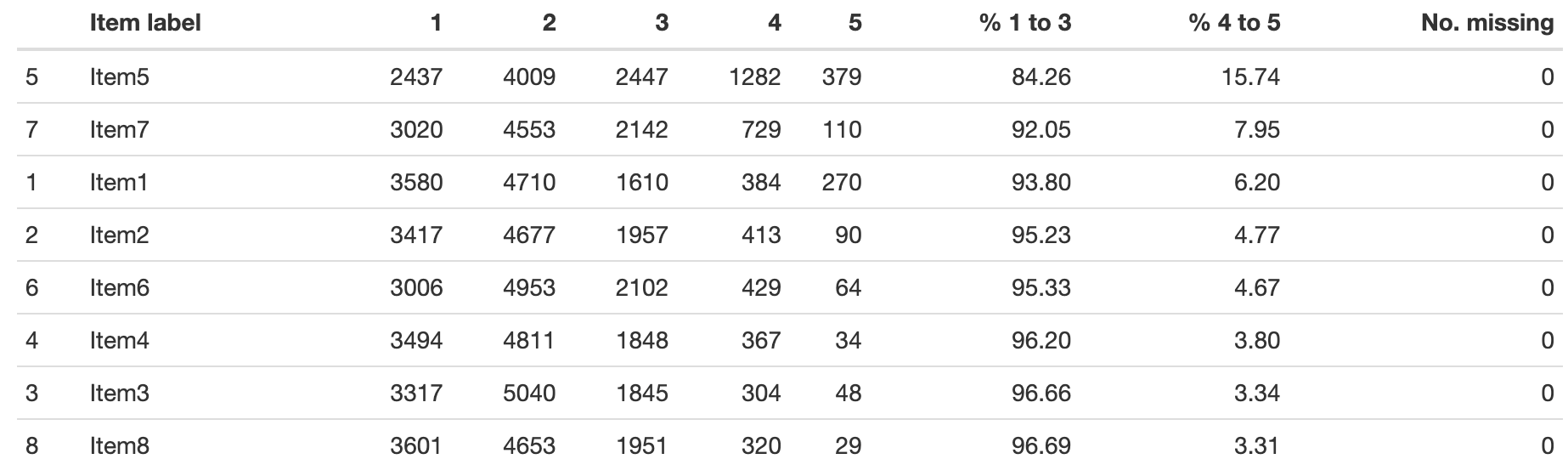

**Table B2. Descriptive statistics for individual items**

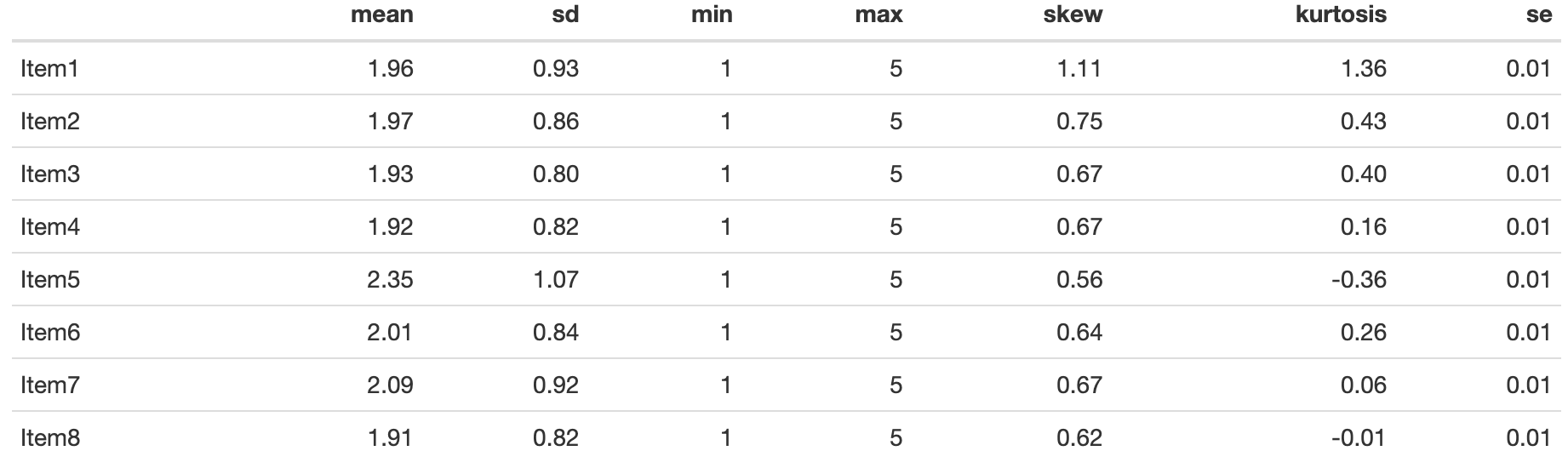

**Table B3. Reliability indices for all scales**

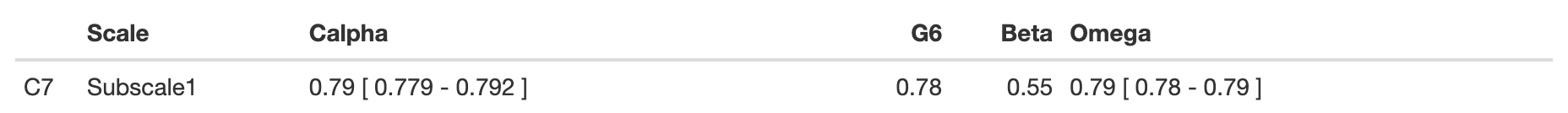

**Table B4. Reliability coefficients with each item removed**

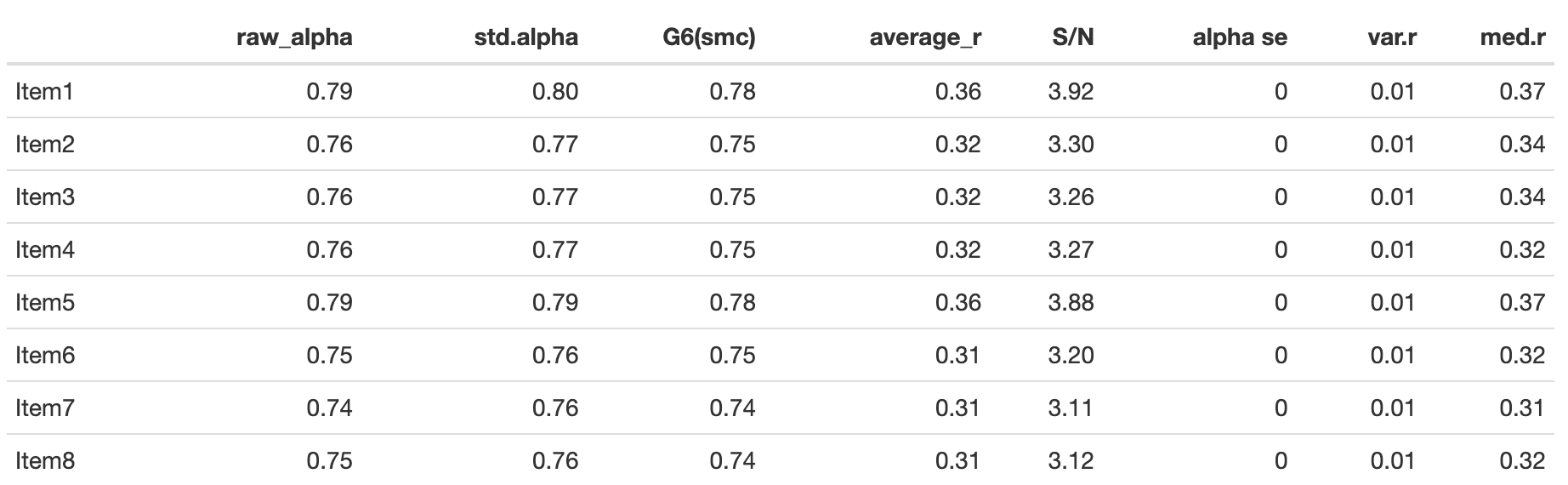

**Table B5: Descriptive statistics for total scale scores**

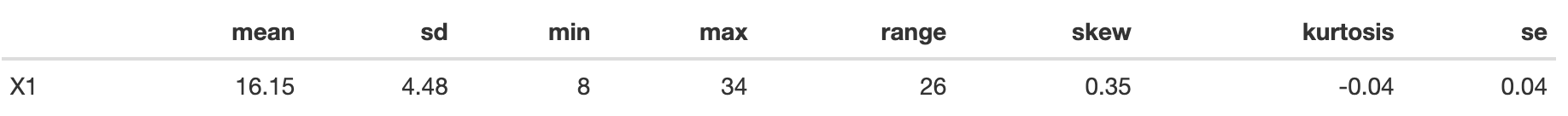

**Figure B1: Bar plots of high score response frequencies**

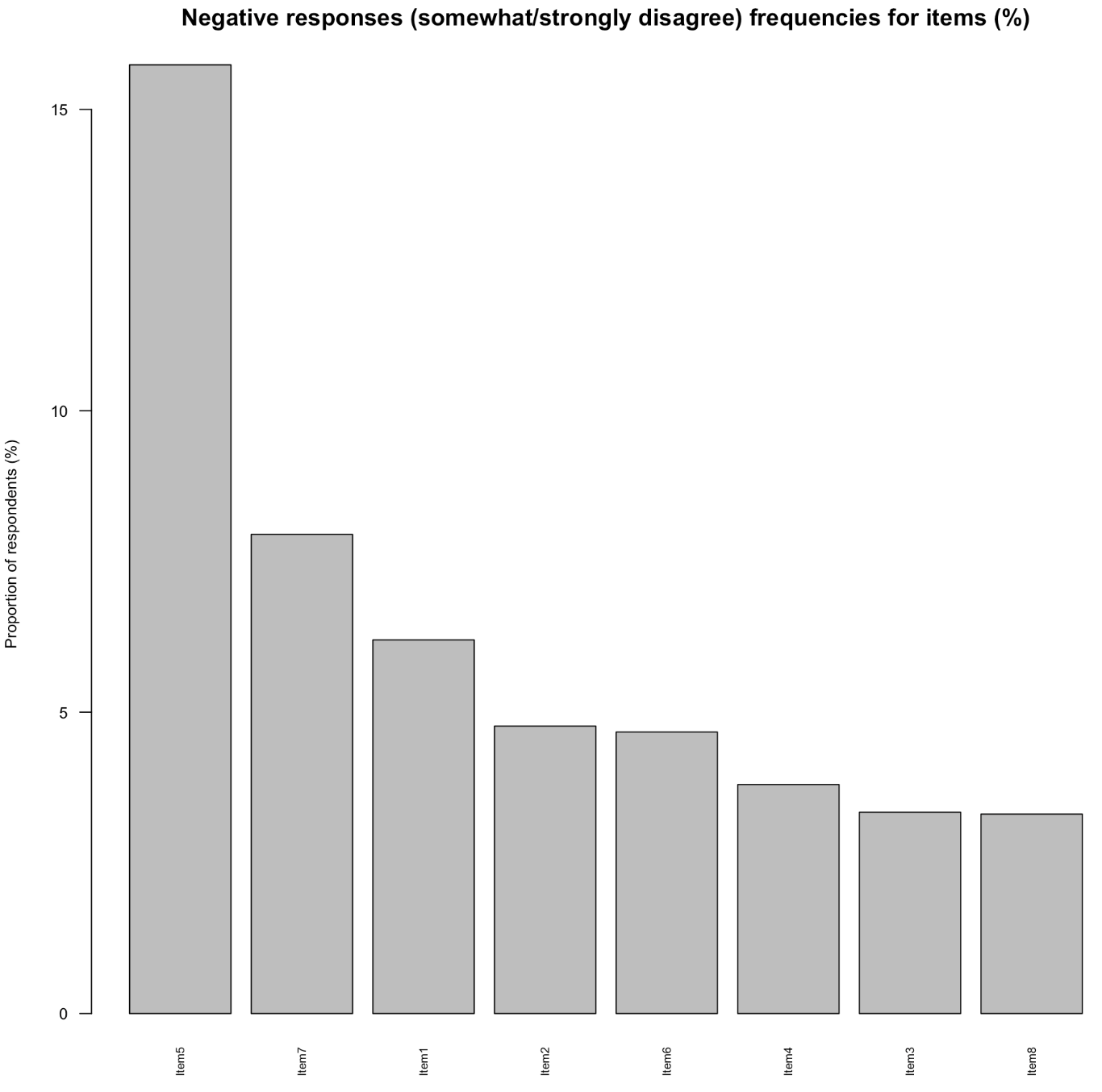

**Figure B2: Bar plots of response frequencies**

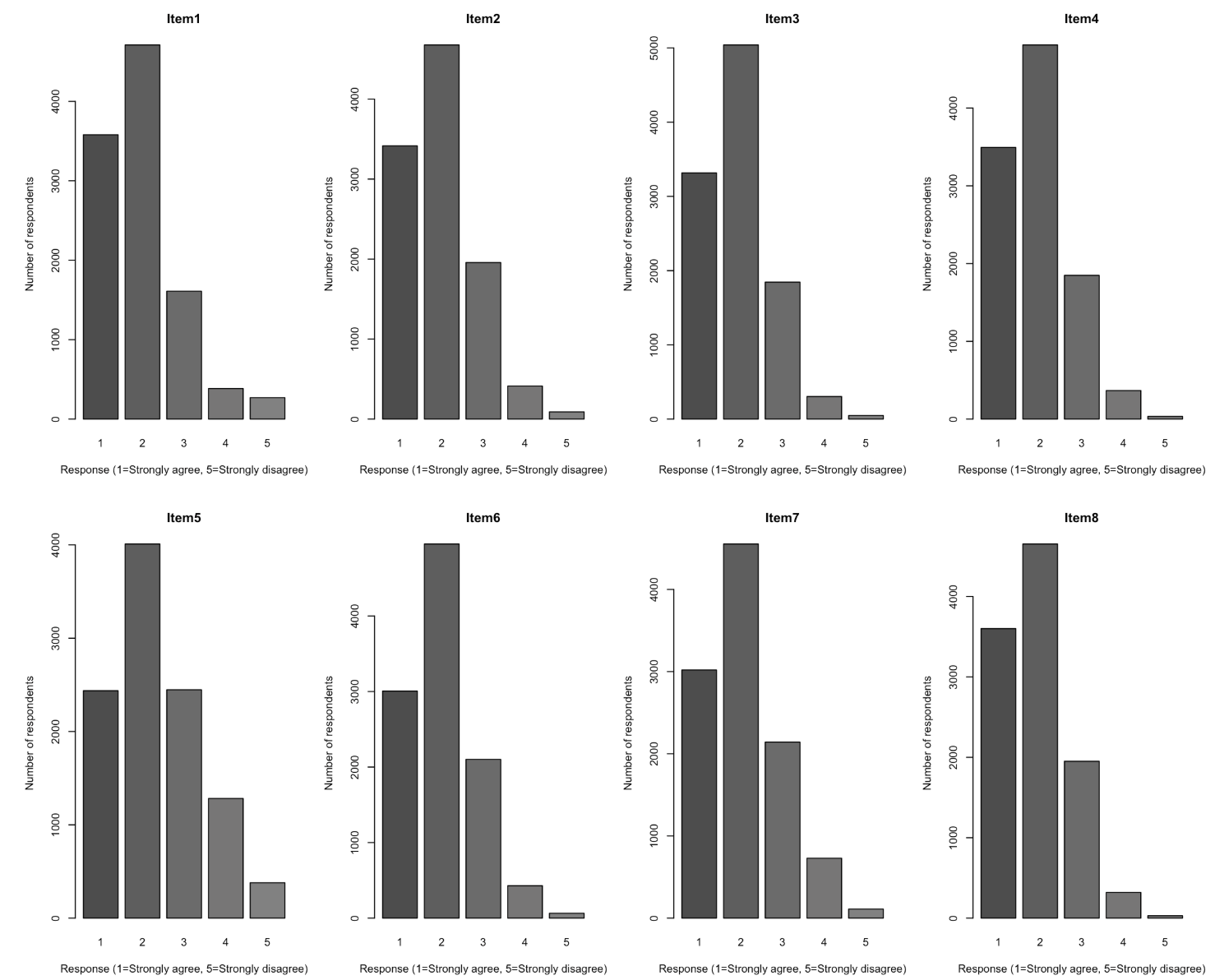

**Figure B3: Heatmap of Spearman correlations among item scores**

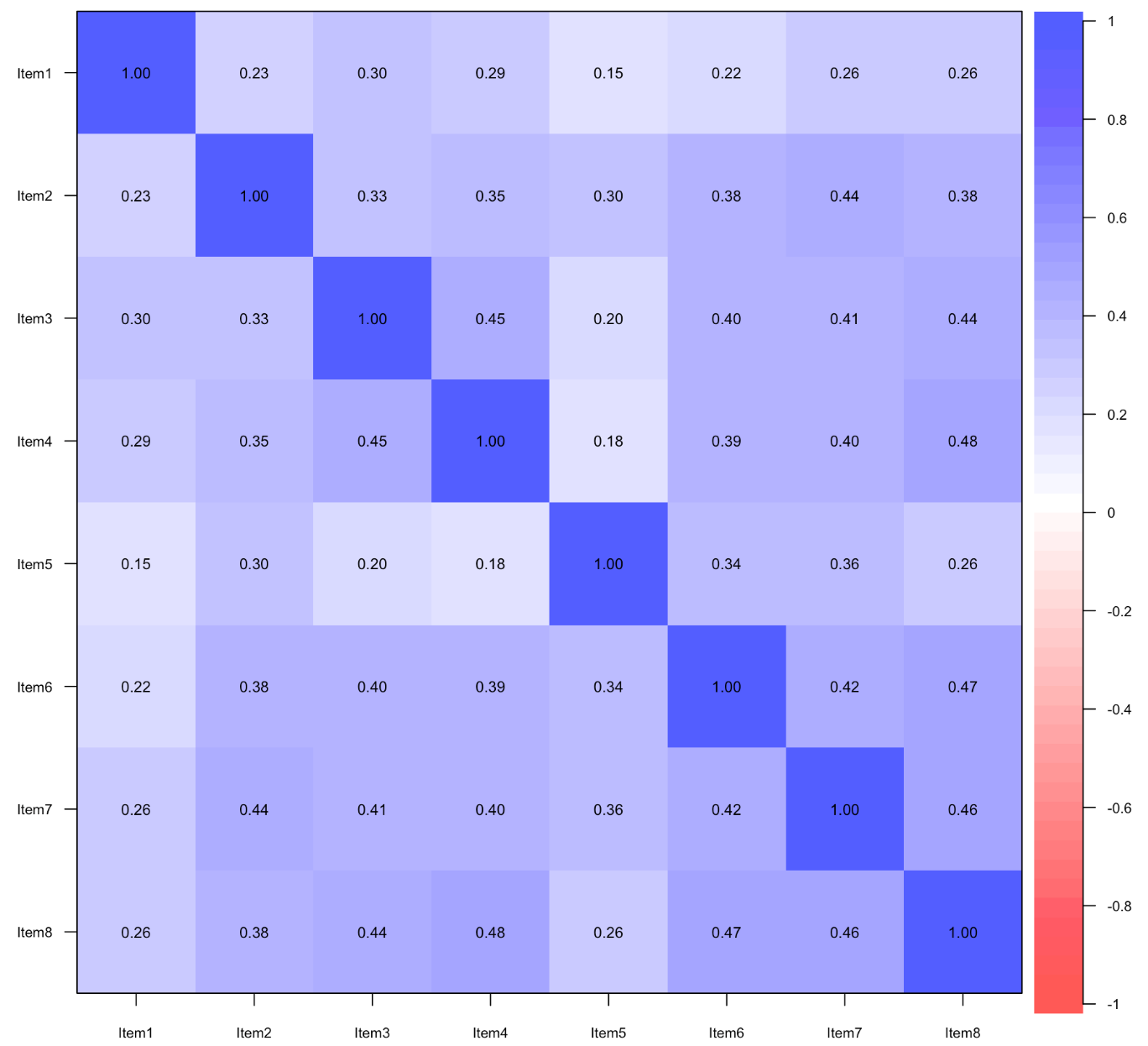

**Figure B4: Identification of multivariate outliers in item set (Q–Q plot)**

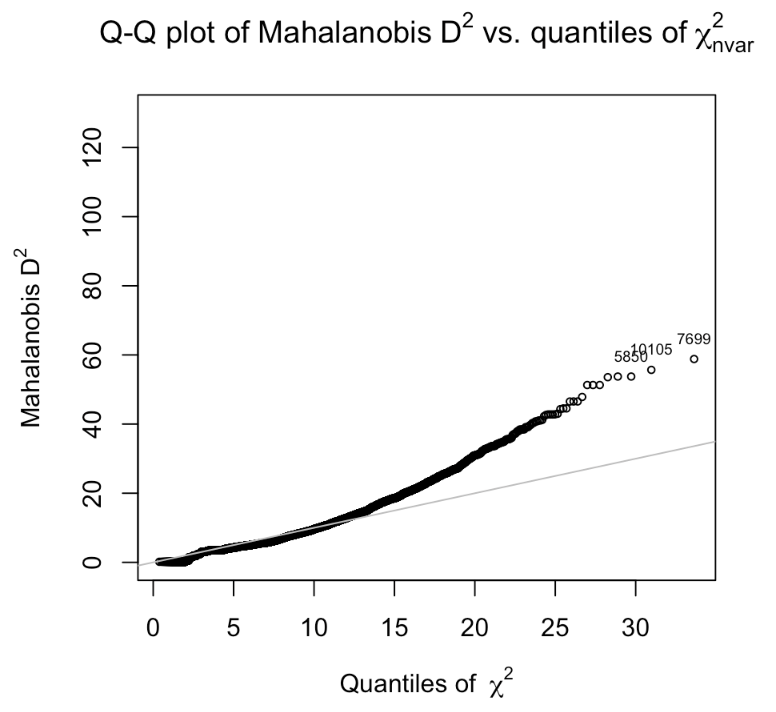

**Figure B5: Identification of multivariate outliers in item set (histogram)**

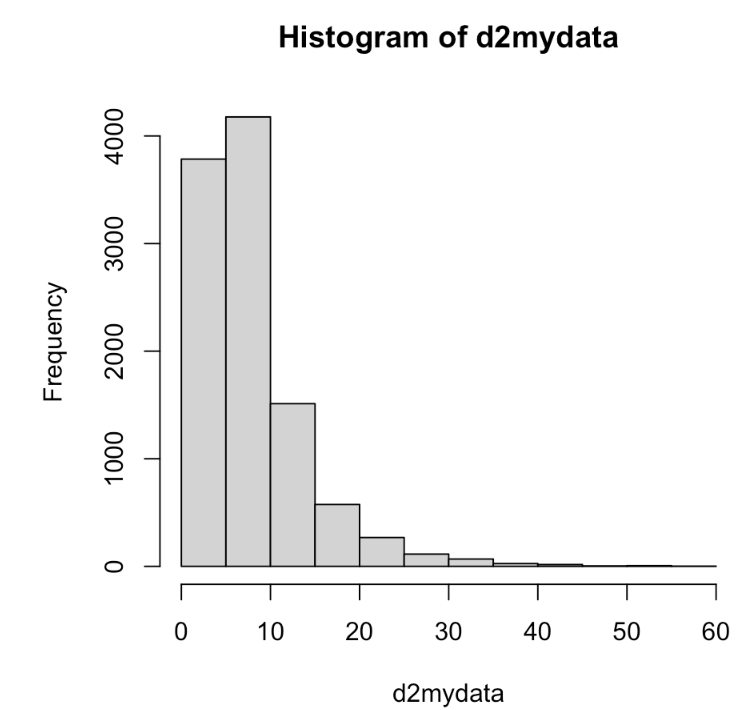

**Figure B6: Parallel analysis scree plot for factor extraction**

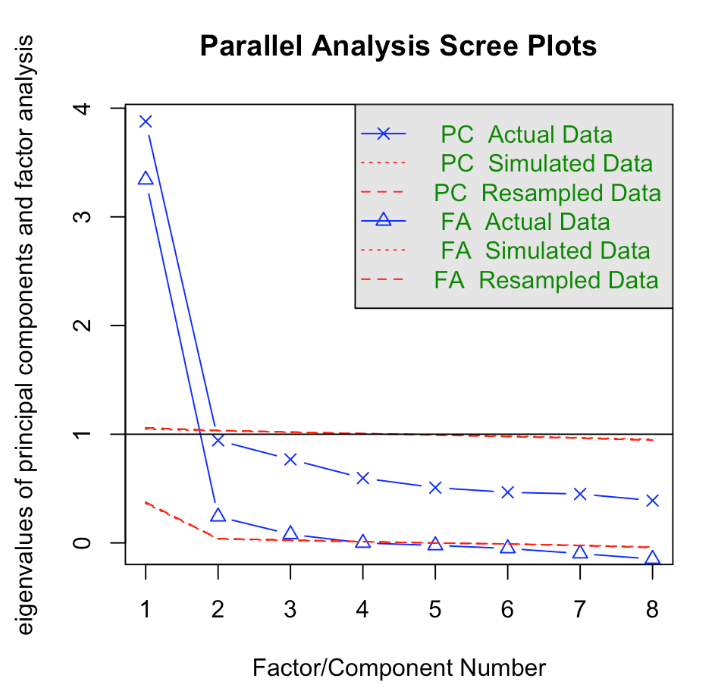

**Figure B7: Very Simple Structure (VSS) plot including outliers**

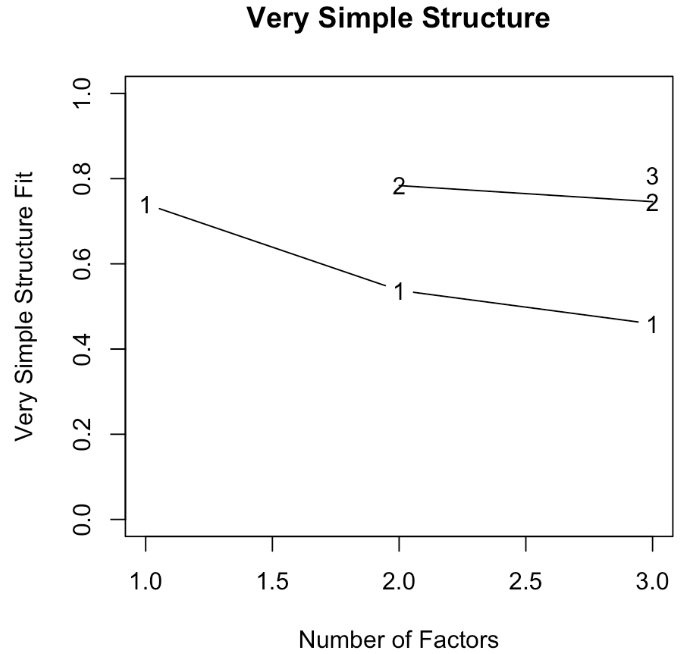

**Figure B8: Very Simple Structure (VSS) plot excluding outliers**

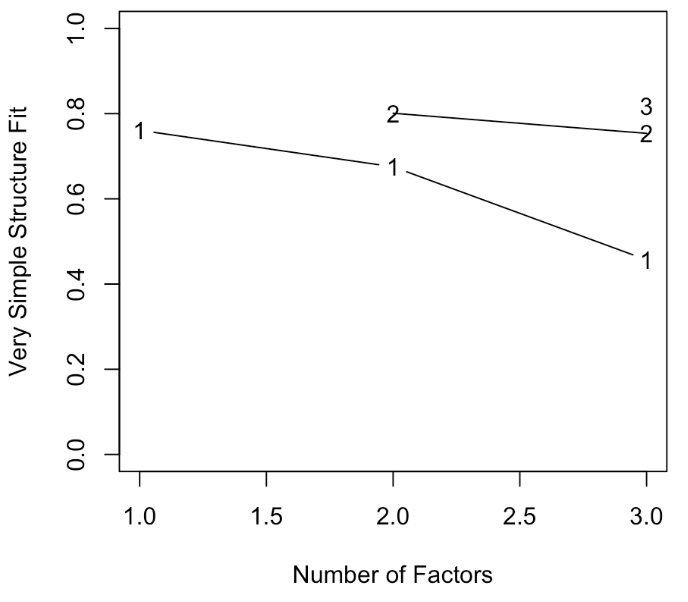

**Figure B9: One-factor exploratory factor analysis (principal axis factoring), including outliers**

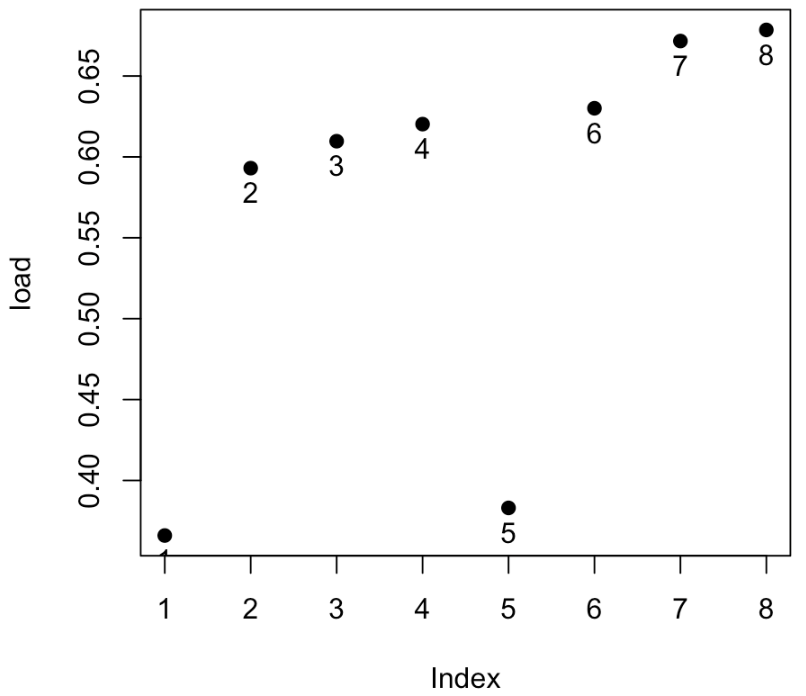

**Figure B10: One-factor exploratory factor analysis (principal axis factoring), excluding outliers**

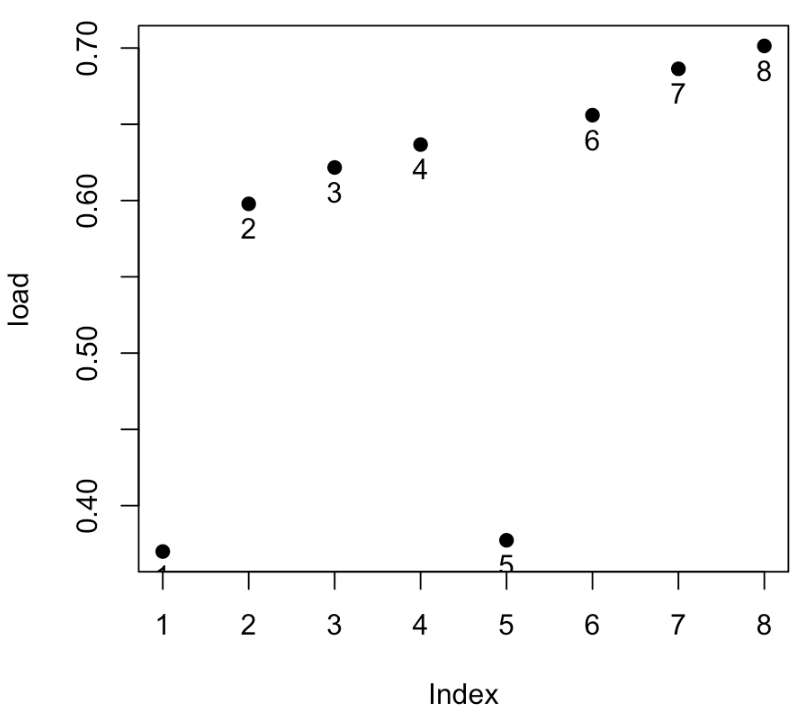

**Figure B11: Hierarchical cluster analysis of scale items (ICLUST), including outliers**

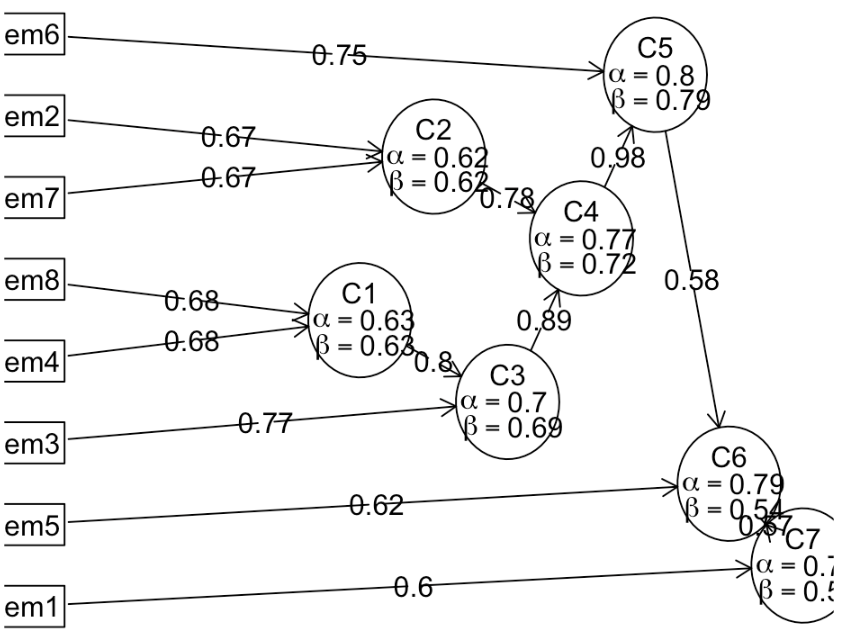

**Figure B12: Hierarchical cluster analysis of scale items (ICLUST), excluding outliers**

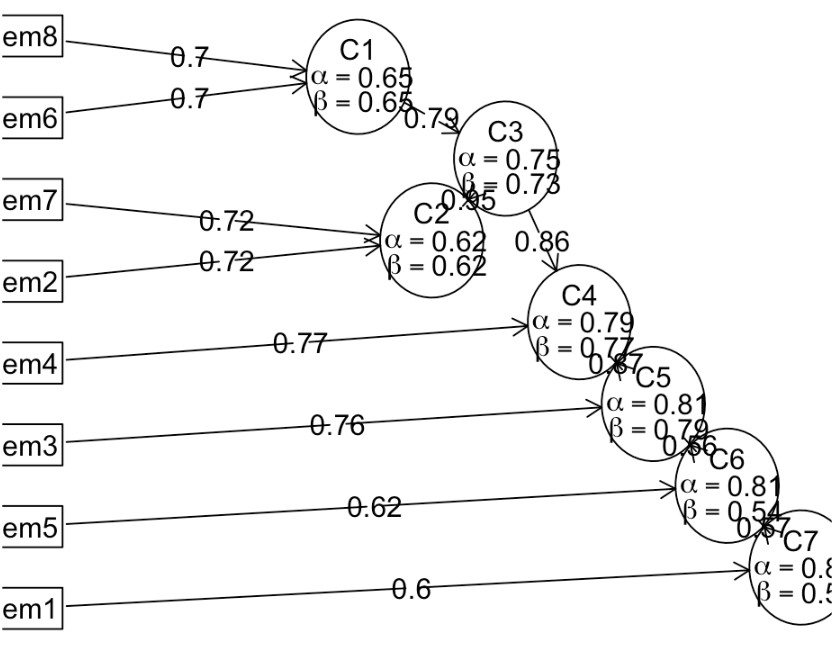

**Figure B13: One-factor confirmatory factor analysis model, including outliers**

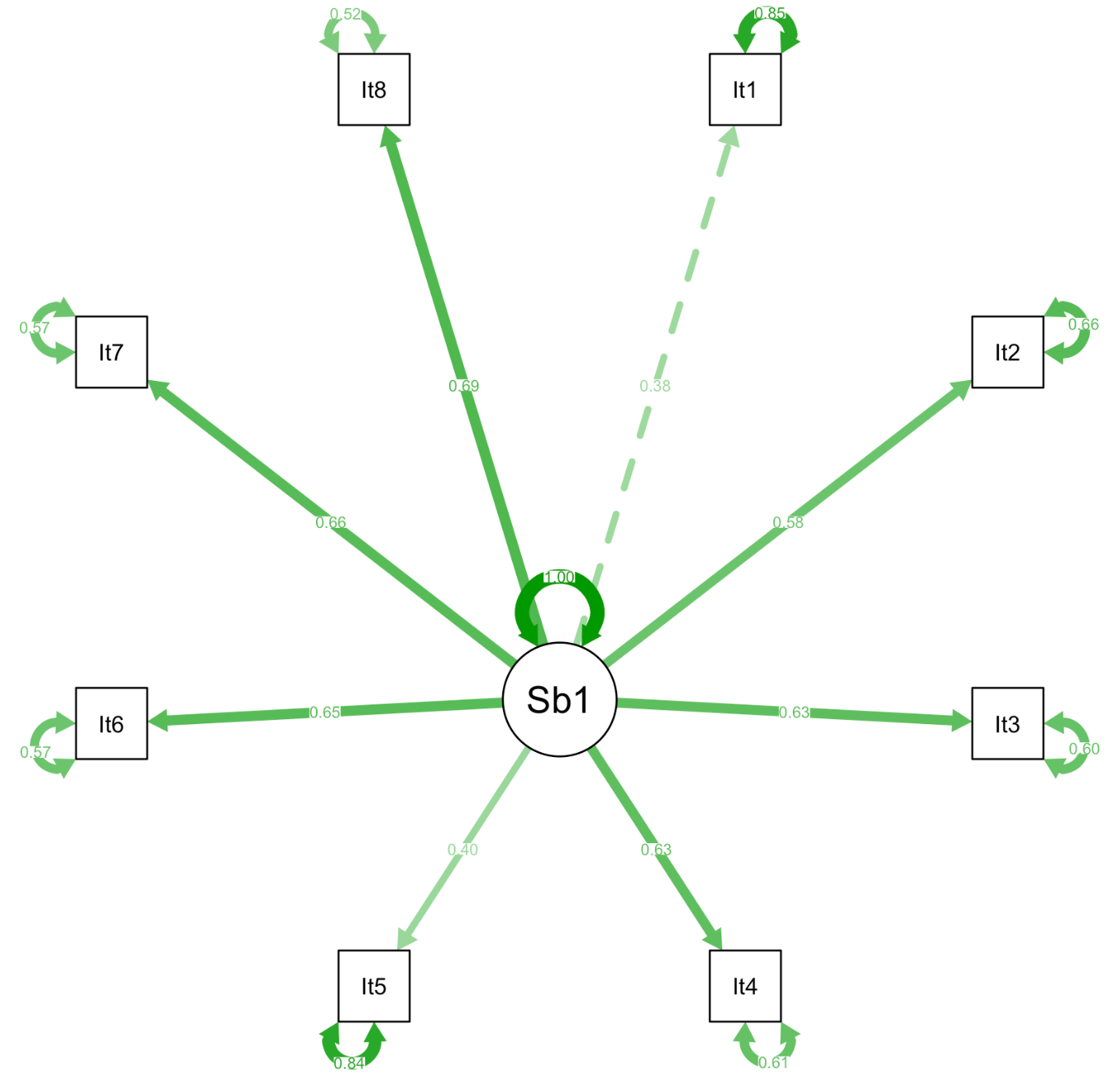

**Figure B14: One-factor confirmatory factor analysis model, excluding outliers**

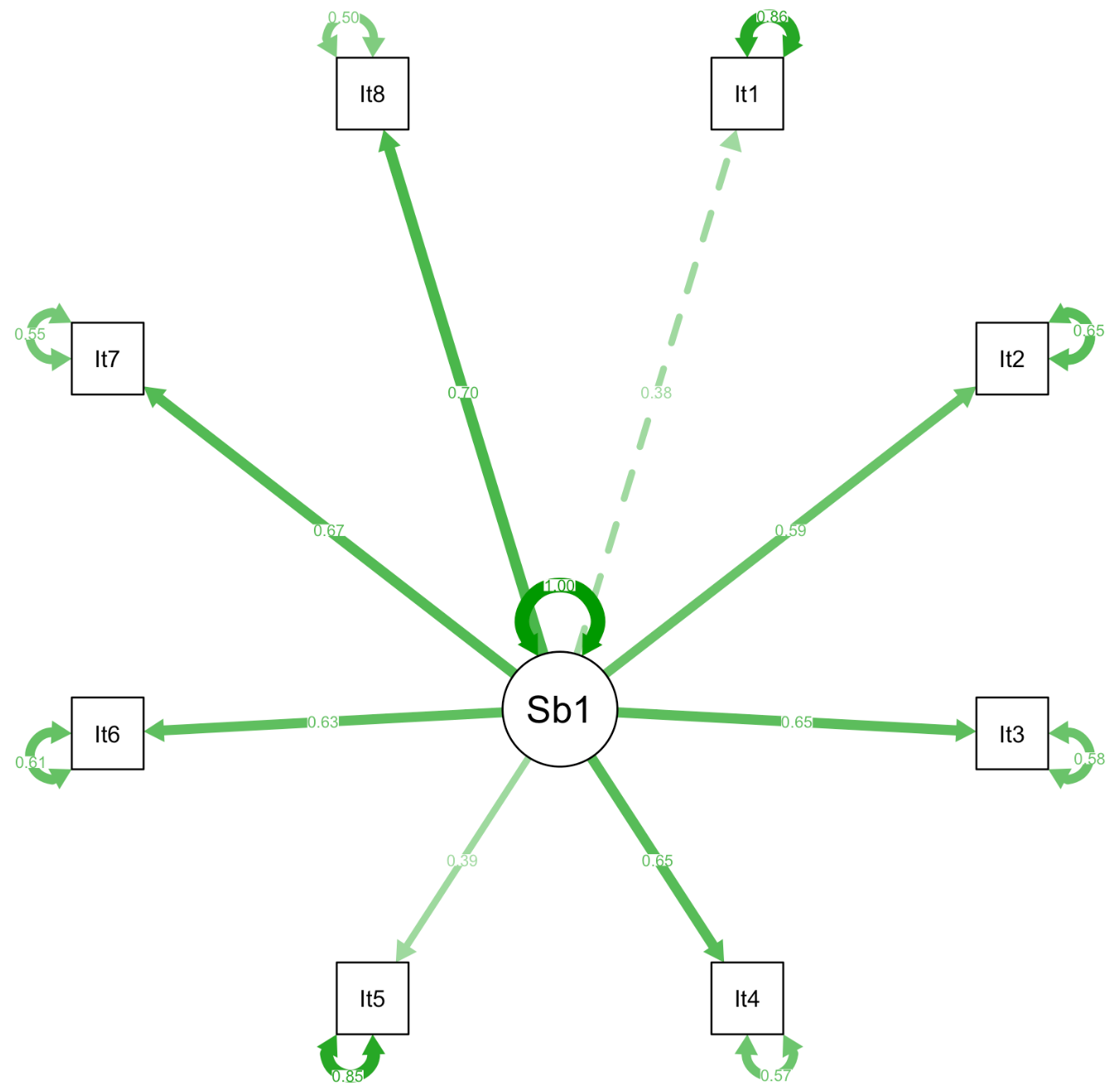

**Figure B15: Distribution of total scale scores**

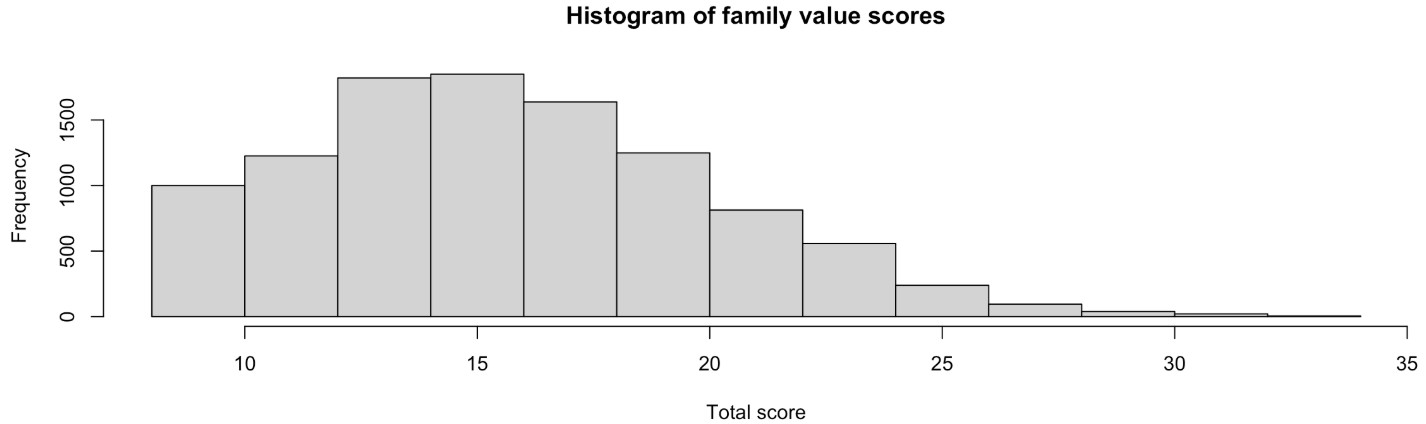
