## Appendix C for "Validation of a New Family Values Scale Among Older Chinese Adults"

**Appendix C: Factor structure of the Items**

**Appendix C1: Correlations between items**

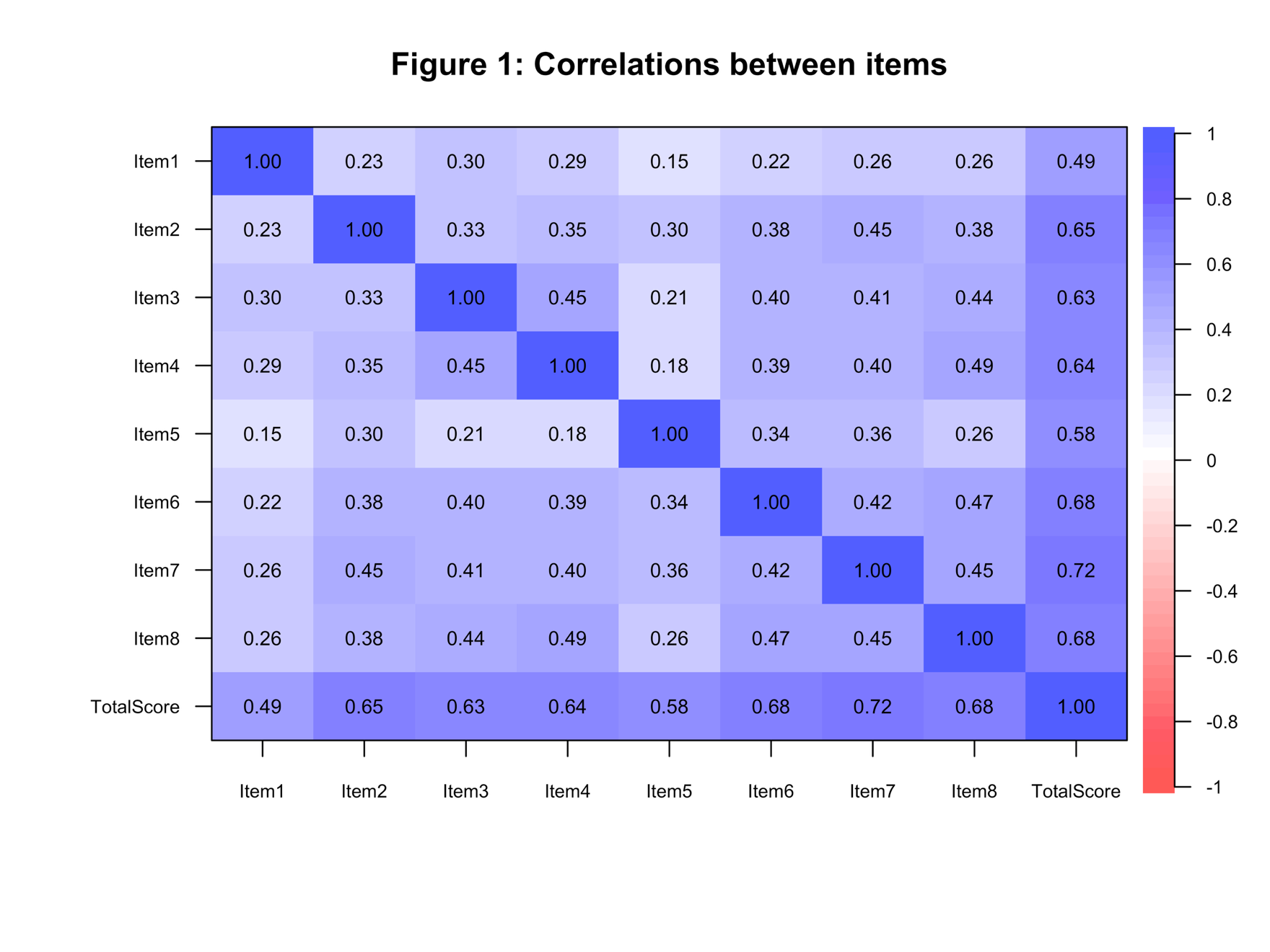

**Appendix C2: Parallel analysis scree plots**

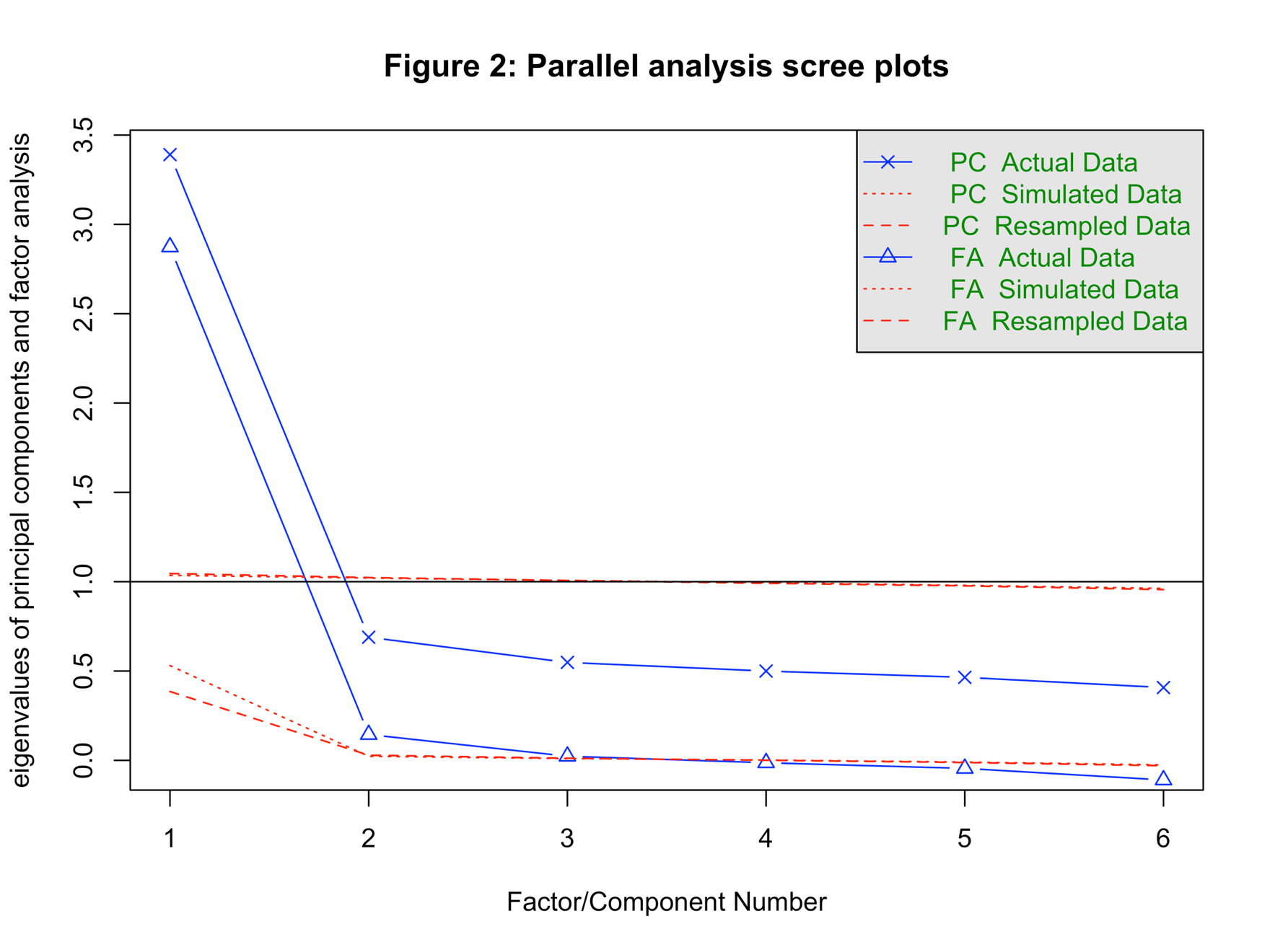

**Appendix C3: Factor loadings of each items**

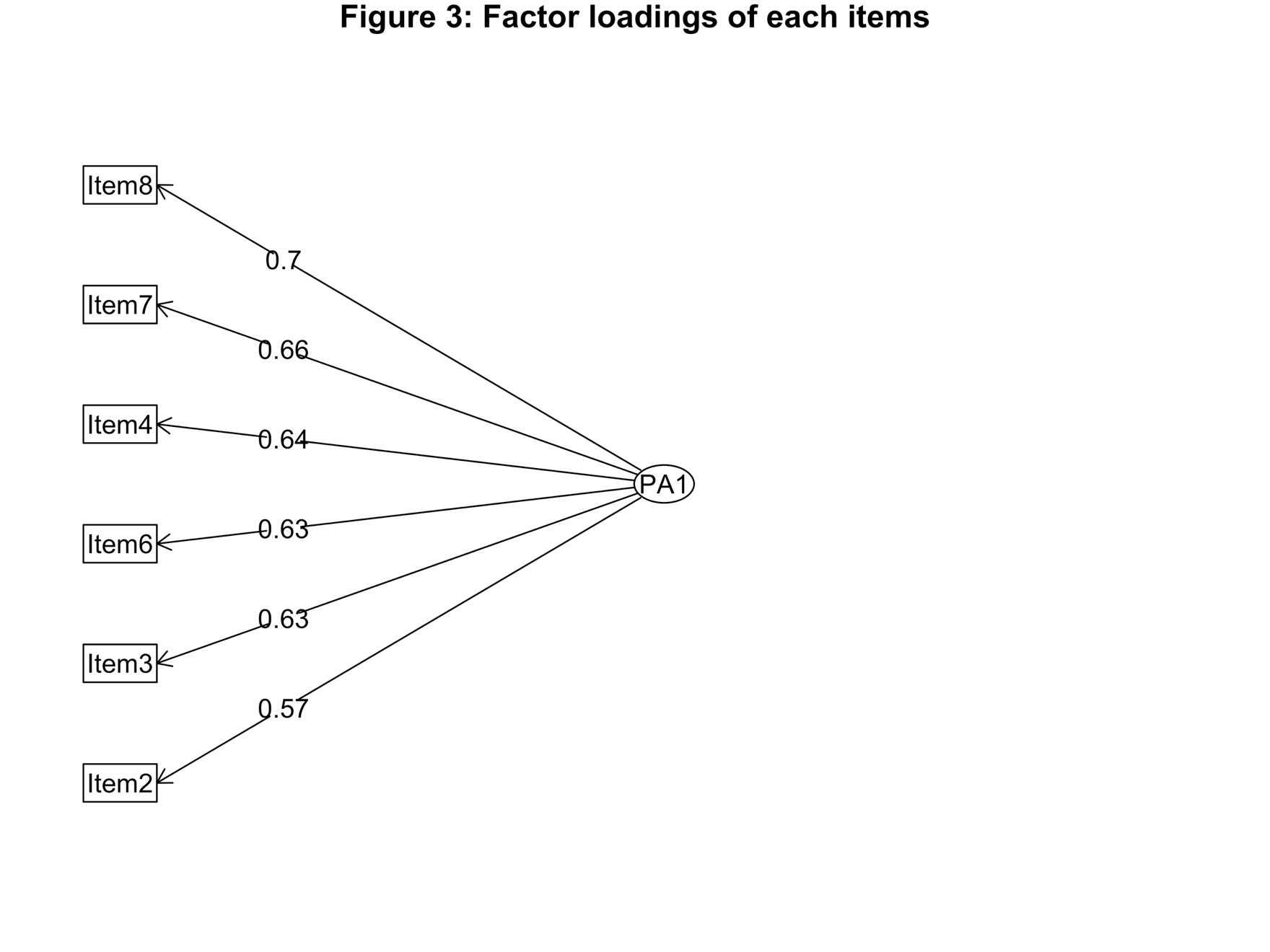
