## Appendix D for "Validation of a New Family Values Scale Among Older Chinese Adults"

**Appendix D: Measurement Invariance Tests**

| **Model** | **CFI** | **RMSEA** | **ΔCFI** | **ΔRMSEA** |
| --- | --- | --- | --- | --- |
| ***Sex (Male: n =*** ***5,736, Female: n = 5,682)*** | | | | |
| Model 1: Configural invariance | 0.985 | 0.052 | NA | NA |
| Model 2: Metric invariance | 0.985 | 0.046 | <0.001 | -0.006 |
| Model 3: Scalar invariance | 0.985 | 0.042 | <0.001 | -0.004 |
| ***Education (Primary school or lower: n = 7,672, Middle school or higher: n = 3,746)*** | | | | |
| Model 1: Configural invariance | 0.983 | 0.055 | NA | NA |
| Model 2: Metric invariance | 0.983 | 0.048 | <0.001 | -0.006 |
| Model 3: Scalar invariance | 0.983 | 0.044 | <0.001 | -0.004 |
| ***Locality (Urban: n = 6,587, Rural: n = 4,831)*** | | | | |
| Model 1: Configural invariance | 0.983 | 0.055 | NA | NA |
| Model 2: Metric invariance | 0.982 | 0.050 | -0.001 | -0.005 |
| Model 3: Scalar invariance | 0.983 | 0.045 | <0.001 | -0.005 |
| ***Geographic regions (Eastern China: n =*** ***5,676, Central China: n = 2,910, Western China: n = 2,832)*** | | | | |
| Model 1: Configural invariance | 0.980 | 0.059 | NA | NA |
| Model 2: Metric invariance | 0.978 | 0.052 | -0.002 | -0.006 |
| Model 3: Scalar invariance | 0.971 | 0.053 | -0.006 | <0.001 |
| ***ADL dependency (Yes: n = 1,786, No: n = 9,632)*** | | | | |
| Model 1: Configural invariance | 0.984 | 0.053 | NA | NA |
| Model 2: Metric invariance | 0.984 | 0.047 | <0.001 | -0.005 |
| Model 3: Scalar invariance | 0.984 | 0.043 | <0.001 | -0.004 |

CFI: Comparative Fit Index.

RMSEA: Root Mean Square Error of Approximation.

ΔCFI, ΔRMSEA: Change in CFI and RMSEA.
